# Epidemiological dynamics of enterovirus D68 in the US: implications for acute flaccid myelitis

**DOI:** 10.1101/2020.07.23.20069468

**Authors:** Sang Woo Park, Jeremy Farrar, Kevin Messacar, Lindsay Meyers, Margarita Pons-Salort, Bryan T. Grenfell

## Abstract

The lack of active surveillance for enterovirus D68 (EV-D68) in the US has hampered the ability to assess the relationship with predominantly biennial epidemics of acute flaccid myelitis (AFM), a rare but serious neurological condition. Using novel surveillance data from the BioFire^®^ Syndromic Trends (Trend) epidemiology network, we characterize the epidemiological dynamics of EV-D68 and demonstrate strong spatiotemporal association with AFM. Although the recent dominant biennial cycles of EV-D68 dynamics may not be stable, we show that a major EV-D68 epidemic, and hence an AFM outbreak, would still be possible in 2020 under normal epidemiological conditions. Significant social distancing due to the ongoing COVID-19 pandemic could reduce the size of an EV-D68 epidemic in 2020, illustrating the potential broader epidemiological impact of the pandemic.

**NOTE:** This preprint reports new research that has not been certified by peer review and should not be used to guide clinical practice.

---

Since 2012, cases of “polio-like” neurological syndromes of uncertain etiology have been reported in the US (*1, 2*) and other countries (*3*). These syndromes, now referred to as acute flaccid myelitis (AFM), are characterized by acute onset of flaccid limb weakness with lesions in spinal cord grey matter (*4*). AFM has affected over 500 individuals in the US alone and exhibits predominantly biennial patterns in the US (*5*) and internationally (*6*). Enterovirus D68 (EV-D68) appears to be the leading cause of AFM, though comprehensive understanding of causality is still incomplete (*7*).

EV-D68 is a nonpolio enterovirus, previously associated with primarily respiratory symptoms (*8*). It was first discovered in 1962 (*9*) and was sporadically reported until 2014, when a large EV-D68 outbreak associated with severe respiratory illnesses occurred in North America; millions of individuals were likely to have been infected in the US during this outbreak, even though only 1,153 cases were confirmed in 49 states (*10*). Since then, EV-D68 outbreaks have reoccurred every other year (2014, 2016, and 2018) in the US, coinciding with major AFM outbreaks (*11–13*). Understanding the epidemiological dynamics of EV-D68 is critical for assessing its causal relationship with AFM and potentially predicting the course of AFM outbreaks.

Nonpolio enterovirus surveillance in the US currently relies on passive reporting through the National Enterovirus Surveillance System (NESS) (*14*). A recent analysis of the NESS data revealed a large geographical variation in the seasonality of nonpolio enteroviruses and the roles of climatic variables in driving seasonal patterns (*15*). However, passive surveillance has several limitations. As enterovirus reports are mostly outbreak-driven, the data are sparse and do not reflect endemic dynamics of enteroviruses; thus only 26 cases of EV-D68 infection were reported between 1970 and 2005 (*14*). The current gap in EV-D68 (as well as other nonpolio enterovirus) surveillance limits our ability to investigate epidemiological dynamics of circulation and association with AFM.

Here, we study EV-D68 dynamics using novel surveillance data collected from the BioFire^®^ Syndromic Trends (Trend) network (*16*). BioFire Trend provides near real-time surveillance based on deidentified clinical test results using the BioFire^®^ System (*16*). EV-D68 cases are then predicted using a Pathogen Extended Resolution (PER) algorithm (see (*16*) for details, and Materials and Methods S1.1 for a summary) from the samples that tested positive for rhinovirus/enterovirus (RV/EV) from the BioFire^®^ FilmArray^®^ Respiratory (RP) Panel results (*17*). We analyze PER-predicted EV-D68 cases across 24 states, consisting of 55 sites, in the US between January 2014 and September 2019. We aggregated the data into 14 regions by combining adjacent states such that each region has at least 3 contributing sites to meet the confidentiality requirements. Separate permission was obtained from the sites in Missouri. This unique data set allows us to characterize detailed spatiotemporal patterns of EV-D68 circulation, which otherwise would not have been possible.

Fig. 1A presents time series of weekly proportion of PER-predicted EV-D68 cases (n=9,913) from RV/EV samples (n=165,036) in the US and in 5 regions with the biggest number of RV/EV samples during the study period: New York (NY; n=37,430), Ohio (OH; n=22,448), Missouri (MO; n=11,706), the Utah and Colorado region (UT+CO; n=20,880), and the Florida, Georgia, and South Carolina region (FL+GA+SC; n=21,273). Time series of weekly proportion (as well as number) of PER-predicted EV-D68 cases and weekly number of RV/EV cases for all 14 regions are presented in Supplementary Materials Fig. S1–S3. These time series demonstrate a clear overall biennial pattern in the US, as well as a considerable variation in the PER-predicted EV-D68 dynamics across different regions (Fig. 1A; Supplementary Materials, Fig. S1–S2). For example, region-level PER-predicted EV-D68 dynamics exhibit regular biennial (NY and UT+CO), multiennial (MO and OH), and irregular dynamics (FL+GA+SC). These observations are consistent with the results of wavelet analyses (Supplementary Materials, Fig. S4–S9; Materials and Methods S1.4). The absence of a major outbreak in Ohio, and possibly Missouri, in 2016 (Fig. 1A; cf. (*18*)) is particularly surprising, given the apparent biennial pattern in the US. Overall, the outbreaks in 2016 appear to be less synchronized, compared to the outbreaks in 2014 and 2018 (Supplementary Materials, Fig. S10).

**Figure 1:**
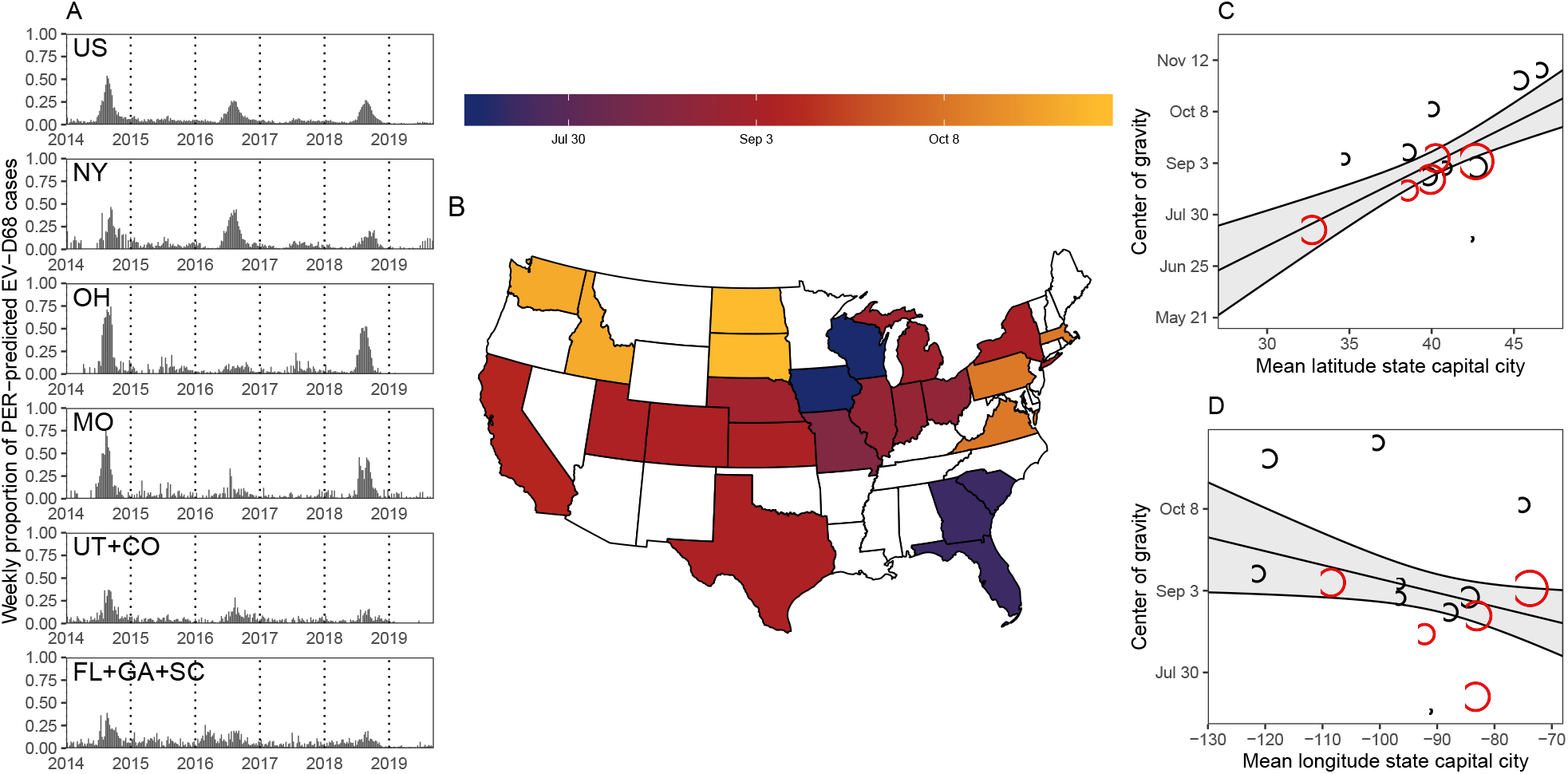
Spatiotemporal dynamics of PER-predicted EV-D68 cases across 24 states (2014– 2019). (A) Weekly proportion of PER-predicted EV-D68 cases. A clear biennial pattern can be observed in the US time series, but local patterns vary. (B) Map of center of gravity of PER-predicted EV-D68 cases. (C) Latitudinal and (D) longitudinal variation in the center of gravity across 14 regions. Red circles represent 5 regions in panel A. Black circles represent the remaining 9 regions. Areas of the circles (and a triangle) are proportional to the number of RV/EV samples. Solid lines represent the estimated regression lines and the associated 95% CIs from the bivariate linear regression; since the bivariate regression depend on two continuous variables, the estimated univariate relationships are plotted by fixing the remaining covariate at its average values.

We characterize the geographical variation in the seasonal patterns of PER-predicted EV-D68 dynamics by calculating the center of gravity (i.e., mean timing of cases (*19*)) for each region (Fig. 1B; Materials and Methods S1.5). The centers of gravity vary from July to November. A bivariate regression analysis of the centers of gravity, using latitude and longitude of the mean state capitals in each region as covariates, suggests a significant, latitudinal increase (0.18 months per degree; 95% CI: 0.11 – 0.26; *p* < 0.001; Fig. 1C) and latitudinal decrease in mean timing of cases (−0.020 months per degree; 95% CI: -0.038 – -0.002; *p* < 0.05; Fig. 1D).

These patterns are consistent with the observed patterns from the analyses of other nonpolio enteroviruses and poliomyelitis based on the NESS data and are likely driven by seasonal climatic trends (*15*).

## Spatiotemporal association between EV-D68 and AFM

A visual comparison of monthly time series of the number of confirmed AFM cases and the proportion of PER-predicted EV-D68 cases at the national level suggests a temporal association, albeit with minor differences in their patterns (Fig. 2A). Notably, the peaks of AFM epidemics occur in September, which is one month after the peaks of PER-predicted EV-D68 epidemics (Fig. 2A). The pairwise cross-correlation between the two time series supports this observation – the strongest correlation occurs at a one-month lag (Fig. 2B). This correlation is significant (*r* = 0.7; *p* < 0.001; Fig. 2C).

**Figure 2:**
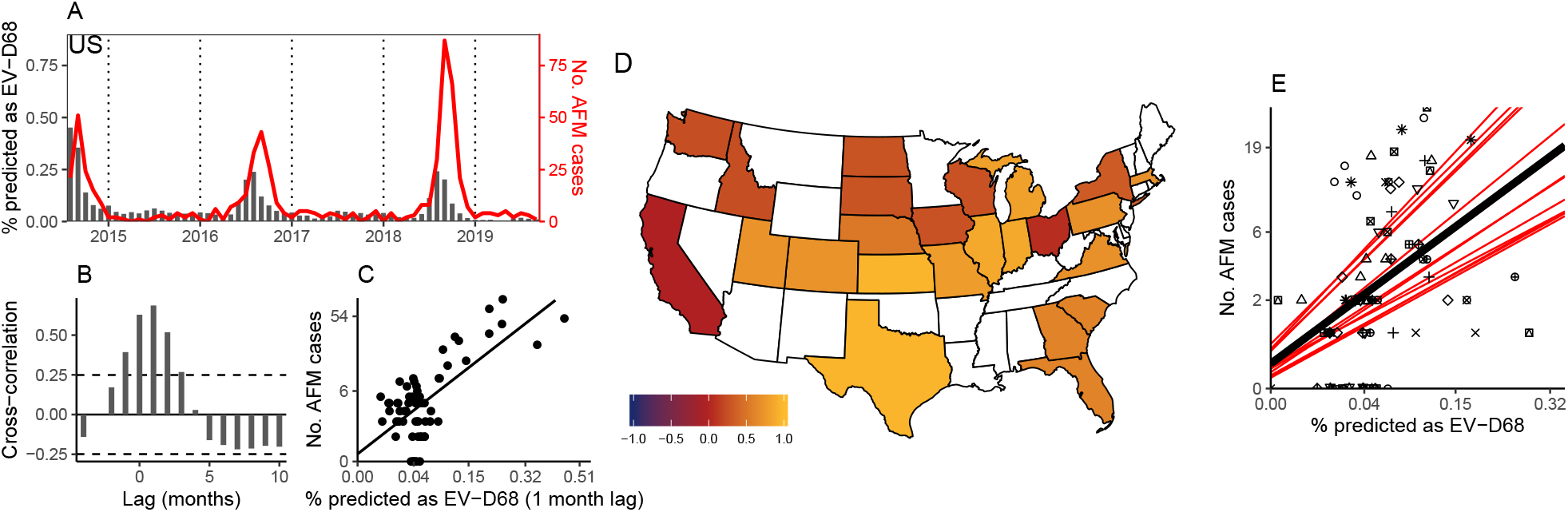
Comparisons of PER-predicted EV-D68 cases and reported AFM cases. (A) Monthly time series of proportion of PER-predicted EV-D68 cases (black bars) and confirmed AFM cases (red lines) in the US. AFM reports are not available prior to August 2014. (B) Pairwise cross-correlation plot between the two monthly time series at the national level using all available EV-D68 and AFM data. (C) Comparison between the monthly proportion of PER-predicted EV-D68 cases at a one month lag and the monthly number of AFM cases at the national level. The solid line is estimated from a linear regression. (D) Map of correlations between annual proportion of PER-predicted EV-D68 cases and the number of AFM cases between 2014–2019 showing a positive association between the two. (E) Comparison between the annual proportion of PER-predicted EV-D68 cases and the annual number of AFM cases across 14 regions (represented by different shapes for each region). Black line represents the estimated fixed effects from a linear mixed model with random slopes and intercepts. Red lines represents the estimated line for each region from a linear mixed model. All correlations are measured after stabilizing time series variances (Materials and Methods S1.3). The axes in Panel C and E are transformed to reflect the variance stabilization.

A one-month lag is longer than the observed delay between onset of prodromal respiratory symptoms and AFM from clinical studies; the estimated median delays range from 2 to 8 days across studies (*20–24*). Our estimated lag is likely to be caused by two approximations. First, the AFM data are reported at a monthly scale, meaning that delays smaller than one month are difficult to assess. Second, there are variations in AFM reporting, which reflects differential awareness and its passive nature (*25*), as well as differential deployment of the BioFire^®^ System (see Supplementary Text S2.1). Given state-level variations in the temporal patterns of EV-D68 circulation (Fig. 1A), comparing the national AFM cases with the PER-predicted EV-D68 cases from 18 states is likely to exaggerate the differences.

The proportions of PER-predicted EV-D68 cases in 2014, 2016 and 2018 remain roughly constant (Fig. 2A), whereas the number of confirmed AFM cases in 2018 (*n* = 238) nearly doubled compared to that in 2014 (*n* = 120). Although this pattern is likely to reflect increased surveillance efforts and awareness of AFM (*25*), other factors could have played a role. For example, phylogenetic analyses of recent EV-D68 outbreaks have demonstrated the spread of different lineages in different years (*26*). Although it remains unclear whether neurotropism is a recently acquired trait (*27, 28*), some studies have suggested that a subset of contemporary EV-D68 strains are more neurovirulent than the prototypical 1962 strain (*27,29*). Future studies may consider whether increased neurovirulence of recent EV-D68 strains is associated with the increased number of AFM cases. Geographically isolated sporadic AFM outbreaks associated with other nonpolio enteroviruses (e.g., enterovirus A71 outbreak in 2018 Colorado (*30*)) could have contributed to the increased number of AFM cases in certain areas.

We find strong (though noisy), positive correlations between the annual number of confirmed AFM cases and the proportion of PER-predicted EV-D68 cases in most regions (Fig. 2D; Supplementary Materials, Fig. S11–13). We use a permutation test to quantify the significance of the average of these correlations (*r* = 0.51; 95% CI: 0.33–0.69; *p* < 0.001). A linear mixed effects analysis with random intercepts and slopes across regions further supports the positive temporal association between AFM and PER-predicted EV-D68 dynamics (*p* < 0.001; Fig. 2E; Supplementary Materials, Fig. S12). Even though there is a large regional variation in the PER-predicted EV-D68 dynamics (Fig. 1; Fig. S1–S2) and the observed AFM dynamics (Supplementary Materials, Fig. S13), we find clear biennial patterns in both across all four census regions (except for the PER-predicted EV-D68 dynamics in the Midwest; Supplementary Materials, Fig. S14). These results indicate that the biennial association between EV-D68 and AFM is robust across the US.

## Epidemiological model fitting

We use an epidemiological model to test whether an interaction between prolonged, serotype-specific immunity and an increase in the susceptible pool via birth (post maternal immunity) can explain the state-level variation in the observed EV-D68 dynamics. Pons-Salort and Grassly showed that the acquisition of serotype-specific immunity can explain complex dynamics observed in other nonpolio enterovirus surveillance data from Japan: in general, large outbreaks reduce the number of susceptible individuals significantly and will be followed by a series of small outbreaks until a sufficient number of susceptible individuals have been added to the population through birth. Like them, we use a stochastic Susceptible-Infected-Recovered (SIR) model that assumes homogeneous mixing, seasonally varying transmission rates, and realistic birth and death rates (Supplementary Materials, Fig. S15; Materials and Methods S1.2). We find maximum likelihood estimates (MLE) of model parameters by fitting the model to time series of proportion of PER-predicted EV-D68 cases between January 2014 and December 2017 from the 4 regions that we presented earlier (Fig. 1); we do not fit the model to the Florida, Georgia, and South Carolina region because two epidemic peaks in 2016 come from different states, which cannot be captured by our model. Fitting the model to data from other regions are precluded by the sparsity of PER-predicted EV-D68 cases. We then assess whether our SIR model can accurately predict out-of-sample EV-D68 dynamics by projecting model fits until September 2019. All confidence intervals are calculated by importance sampling (*32*). A detailed explanation of the methods is given in Materials and Methods S1.6–S1.7.

Our simple, mechanistic model can capture the PER-predicted EV-D68 dynamics from 2014 to 2017 in all 4 regions (Fig. 3A). Our model fits also demonstrate that the observed regional variations in EV-D68 dynamics are consistent with model parameter variations; however, some of these variations may be caused by the brevity and crudeness of the time series, as well as the simplicity of the model (see Discussion), and may not reflect real differences among states. All parameter estimates are presented in Supplementary Materials Fig. S16–S20 and Table S2–S5.

**Figure 3:**
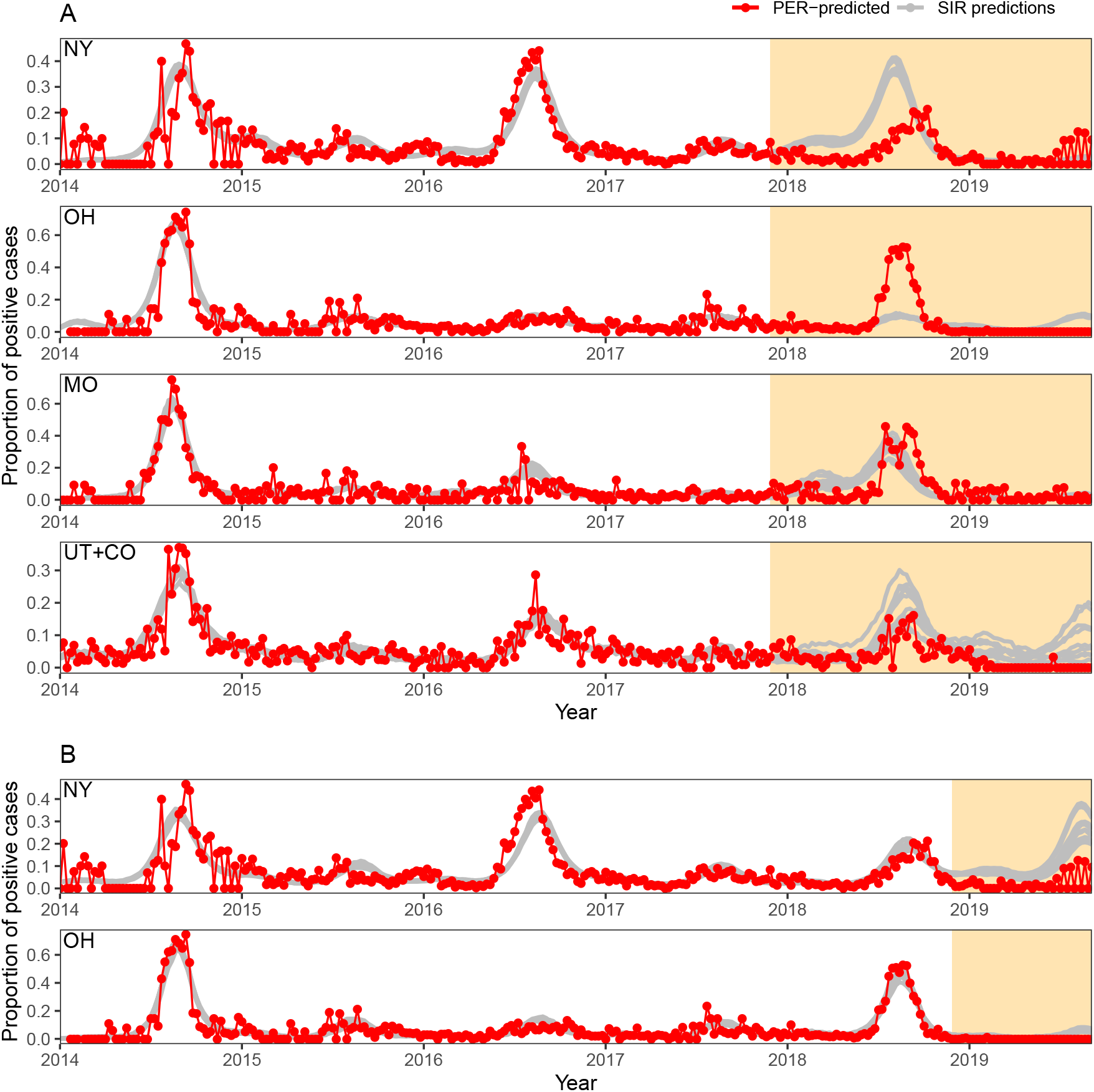
Model fit to data for New York, Ohio, Missouri. (A) A comparison of the PER-predicted (red lines) and model-predicted (grey lines) weekly proportion of positive cases. Predictions were made by simulating forward from 10 particle filtered trajectories. (B) A comparison of the PER-predicted and model-predicted weekly proportion of positive cases, including the outbreak in 2018 into our parameter estimation procedure.

We estimate similar ranges of time-varying effective reproductive number *ℛ*_*t*_ (i.e., the average number of secondary cases caused by an infected individual over the course of an epidemic (*33*)) between 2014 and 2017 for all 4 regions (Supplementary Materials, Fig. S21): New York (0.6–1.4), Ohio (0.3–1.3), Missouri (0.6–1.5), and the Utah and Colorado region (0.8–1.3). However, there is a large variation in the estimates of the basic reproductive number *ℛ*_0_ (i.e., the average number of secondary cases caused by an infected individual in a fully susceptible population): New York (29.5; 95% CI: 26.8–32.7), Ohio (10.7; 95% CI: 8.4–13.0), Missouri (20.6; 95% CI: 17.0–27.7) and the Utah and Colorado region (14.6; 95% CI: 10.9– 18.5). Since epidemiological dynamics of endemic diseases, such as EV-D68, are driven by *ℛ*_*t*_, high estimates of *ℛ*_0_ are likely to be correlated with low estimates of *S*(0). Not explicitly accounting for age structure, metapopulation structure, and heterogeneity in the population is also likely to have affected our estimates (*34*).

Predicting future EV-D68 dynamics from a short time series is difficult. Our model predicts EV-D68 outbreaks in the Utah and Colorado region and in Missouri reasonably well but imperfectly in Ohio and New York. While the model predicts a large outbreak New York in 2018 (Fig. 3A), the observed outbreak in 2018 occurred later in the year and was smaller than the outbreaks in 2014 and 2016. This mismatch suggests that the observed pattern of EV-D68 spread in 2018 New York may be different from the biennial outbreaks observed in 2014 and 2016.

We tested whether our simple model can explain the patterns in 2018 by fitting the same model to the New York and Ohio time series, including the outbreak in 2018. We find that our model can then explain the apparent changes in the EV-D68 dynamics in 2018 reasonably well (Fig. 3B). However, the new model fit does not necessarily provide better predictions for the dynamics in 2019; this lack of fit is partly driven by anomalies in the data (e.g., the PER algorithm predicts almost no EV-D68 cases in Ohio and in the Utah and Colorado region in 2019).

Including the outbreaks from New York and Ohio in 2018 into our model fitting yields much closer estimates of *ℛ*_0_: New York (17.0; 95% CI: 15.9 – 19.7) and Ohio (14.8; 95% CI: 13.8 – 16.5). These changes in the estimated basic reproductive number can suggest a transition in the long-term dynamics to cycles of different periodicity (*35*). While both parameter sets give *qualitatively* similar fits to 2014–2017 time series, the initial parameter estimate provides a *quantitatively* better fit for the first 4 years (a difference of 17 and 21 log-likelihood units for New York and Ohio, respectively); these differences confirm that our initial parameter estimates are well-identified. Given the brevity of the time series, it is not surprising that there are multiple parameter combinations that give qualitatively similar short-term dynamics but different long-term dynamics.

Finally, we explore the long-term stability of the EV-D68 attractors using a bifurcation diagram of the deterministic SIR model (Fig. 4A; Materials and Methods S1.8). We rely on estimated parameters for New York, since it has the best available data for EV-D68. The bifurcation diagram predicts predominantly annual cycles in Ohio (excluding 2018) and the Utah and Colorado region although multiple attractors may coexist (indicated by the fine-grained pattern of colors representing different cycle periods; cf. (*35*)); dynamical transitions due to stochasticity may drive irregular patterns in these regions. Our model predicts a stable, long-term biennial cycle in New York (excluding 2018) and Missouri, but their parameters lie near the edge of a dynamical transition that divides the biennial regime from irregular (New York) and annual (Missouri) regimes. On the other hand, parameter estimates that include the 2018 outbreaks in New York and Ohio are closer together and are within an annual regime, suggesting that the patterns observed in 2014-2017 might have been a transient feature. Although these results suggest that the observed biennial pattern may not be stable in the long term, stochastic simulations show that a moderate to large outbreak, comparable to that in 2014, is possible between August and September 2020 across the ranges of parameters we estimate for New York if epidemiological conditions remain unchanged (Fig. 4B).

**Figure 4:**
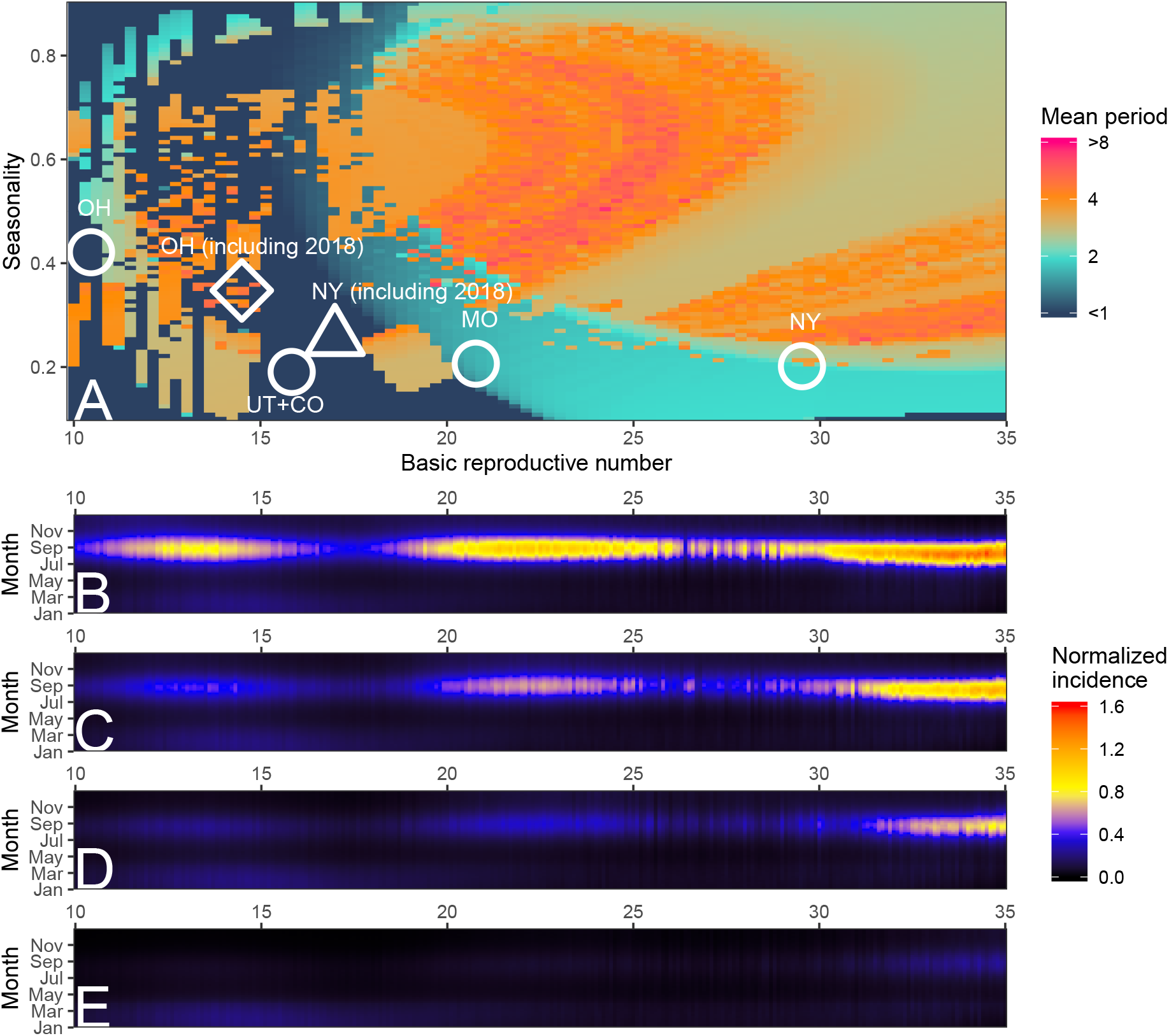
Long- and short-term predictions of EV-D68 dynamics. A. Bifurcation diagram of the deterministic SIR model using parameter estimates for New York but by varying *ℛ*_0_ and the relative amplitude of the seasonal forcing. Colors indicate the mean periodicity of the cycles. Circles represent the original parameter estimates for each state. The triangle and diamond represent the new parameter estimates that include the 2018 outbreak for New York and Ohio, respectively. B–E. Predictions of EV-D68 outbreak in 2020 using stochastic simulations using parameter estimates for New York (B) and assuming 5% (C), 10% (D), and 20% (E) decrease in transmission rates from the 12th week of 2020 until the end of 2020. For each *ℛ*_0_ value, we let *S*(0) = 1.04*/ℛ*_0_ and run 100 stochastic simulations until December 2020. We calculate the normalized incidence by dividing the medians of the model-predicted weekly incidence by the median of the model-predicted peak incidence in 2014.

In practice, massive non-pharmaceutical (social distancing) efforts in response to the on-going COVID-19 pandemic could reduce average contact rates during 2020 (*36*). We explore the potential impact of these actions on EV-D68 dynamics in Fig. 4C–E. If average transmission rate is reduced by more than 10%, a major EV-D68 outbreak is unlikely to occur. Note that we do not explore longer term dynamics, arising from the consequent increase in EV-D68 susceptibility.

## Discussion

Here, we present the first spatiotemporal analysis of EV-D68 dynamics in the US and its population correlation with cases of AFM. Our analysis reveals spatial variation in the patterns of spread of EV-D68. We showed that these variations are correlated with latitude and longitude and can be explained by a simple SIR model at the state level. These observations are consistent with previous studies of enteroviruses and are likely climate-driven (*15, 31*). Finally, our analyses suggest that the predominant medium-term biennial dynamics in the spread of EV-D68 may not necessarily be stable. Given uncertainties in EV-D68 dynamics as well as in behavioral changes due to the COVID-19 pandemic, the occurrence or the size of EV-D68 outbreaks in 2020 remains unclear.

There is currently a lack of data to characterize more detailed epidemiological dynamics of EV-D68. Even though EV-D68 has been circulating for at least 60 years (*9*), our data from the PER algorithm, which predicts the presence of EV-d68, only span the last 6 years. The passive surveillance data is expected to be even sparser. Based on such short time series, it is difficult to assess whether the observed patterns will persist. Nonetheless, our simple model captures the observed dynamics reasonably well and allows us to make useful predictions about the course of the epidemic; these predictions will become firmer as more data becomes available. More broadly, this analysis underlines the power of leveraging novel sources of large-sample surveillance data (*37*). While the BioFire Trend data permit a step change in analysis, surveillance based on constant sampling effort would be much more powerful. Population-representative incidence and serological data by state (as well as internationally) would allow us to further characterize the dynamics of EV-D68 and its association with AFM.

An important criterion for probing the causality between EV-D68 and AFM is their temporality at the individual level (i.e., infection of EV-D68 has to precede the onset of neurological symptoms (*7*)), which is supported by the observed onset of respiratory illnesses (*38*) and the detection of EV-D68 (*39,40*) prior to the onset of AFM. Our cross-scale comparison of EV-D68 and AFM dynamics revealed robust correlations between the timing of EV-D68 and AFM cases, providing additional ecological support for a causal link.

Our dynamic model relied on several simplifying assumptions about the biology of EV-D68. For example, we assumed that the infectious period distribution follows an exponential distribution with a mean of 7 days. Although Pons-Salort and Grassly (*31*) showed that nonpolio enterovirus dynamics in Japan can be explained sufficiently well under the same assumption, assumptions about the shape of the infectious period distribution can affect model dynamics (*41, 42*) as well as *ℛ*_0_ estimates (*43*). More importantly, ignoring potential uncertainties in model parameters (e.g., assuming that the mean infectious period is known) can underestimate the overall uncertainties associated with the conclusion (*44*). We also did not test for other mechanisms (e.g., changes in pathogenicity, antigenicity or transmissibility (*31*) or interactions with other nonpolio enteroviruses (*45*)) that could account for the apparent change in epidemiological dynamics in New York in 2018. Future phylodynamics models (*46*) should explore integrated epidemiological and viral genetic dynamics. We also did not account for age structure, strength of immunity, and spatial coupling between states; these simplifications are likely to have affected our estimates of the basic reproductive number. Predictions about the stability of the biennial cycles should be interpreted with caution as our analysis relies on short time series. Our preliminary analysis of the impact of the coronavirus pandemic is necessarily crude. For instance, SARS-CoV-2 and EV-D68 are likely to have very different mean ages of transmission. Nonetheless, current behavioral changes are profound across ages; significant impact of EV-D68 dynamics seems likely.

Despite the simplicity of the analysis, we can draw robust conclusions about key aspects of EV-D68 dynamics. First, the PER-predicted EV-D68 dynamics are consistent with the importance of transmission-blocking herd immunity. Second, although our estimates of the *ℛ*_0_ are necessarily approximate, they are likely to be relatively high (*>* 10). This is consistent with a low median age of infection for EV-D68 (5 years (*47*)) as well as *ℛ*_0_ estimates for other nonpolio enteroviruses exhibiting biennial cycles in Japan (e.g., *ℛ*_0_ = 16.2 for Coxsackievirus A2 and *ℛ*_0_ = 32.7 Coxsackievirus A4 (*31*)). The successful SIR model fits also imply that vaccination could tentatively control the spread of EV-D68, but herd immunity thresholds are likely to be high (*31*).

Although there is large uncertainty associated with the stability of the observed biennial pattern, we predict that a major EV-D68 outbreak can occur in the near future. Based on the low number of PER-predicted EV-D68 cases in 2019, we would expect the number of susceptible individuals to have increased, enhancing the probability for a large outbreak to occur. Our preliminary analysis indicates that the COVID-19 pandemic response may significantly impact the dynamics of a 2020 EV-D68 outbreak; if social distancing prevents the outbreak from occurring, the susceptible pool may increase even further. There are several such potential impacts of pandemic social distancing on other infections; predicting these effects and testing such models against the actuality will be a powerful test of the effects of contact reduction. We propose that mainly ‘ecological’ interactions, such as between SARS-CoV-2 and EV-D68 (*48*), will generate weaker effects than interactions mediated by acquired immunity with SARS-CoV-2’s close relatives, which affect the susceptible pool (*49*).

Improved real-time surveillance, such as the BioFire Trend system, may serve as an important early warning system for impending outbreaks and improve epidemiological modeling to better understand and predict future patterns of circulation. As the PER algorithm is likely no longer to function after this year due to a product update, future research should therefore prioritize developing specific, and accurate, demographically and geographically representative testing for EV-D68 and its enteroviral relatives. More generally, current events underline the importance in future of refining global pathogen and immunological surveillance against the range of potential threats.

## Data Availability

Data used in this study has been aggregated to further protect both the privacy of the patient for
publication and the identity of the contributing institutions. Access to data used in the study
may be made available upon request.

## Acknowledgments

We thank Nicholas Grassly, the CDC AFM team, and members of the CDC AFM Taskforce for much help and useful discussions. We thank Kirsten St. George and New York State Department of Health for providing the AFM data for the New York state. We thank Colorado Department of Public Health and Environment for providing the AFM data for Colorado. SWP thanks Ben Bolker and Jonathan Dushoff for their statistical advice.

## Data availability

Data used in this study has been aggregated to further protect both the privacy of the patient for publication and the identity of the contributing institutions. Access to data used in the study may be made available upon request.

## Funding

KM receives grant support from NIAID (Grant Number K23AI128069). MP-S is a Sir Henry Dale Fellow jointly funded by the Wellcome Trust and the Royal Society (Grant Number 216427/Z/19/Z).

## S1 Materials and Methods

### S1.1 Epidemiological data

The Pathogen Extended Resolution (PER) algorithm, designed to distinguish EV-D68 from other rhinoviruses/enteroviruses (RV/EV), was developed using data from the BioFire^®^ FilmArray^®^ Respiratory (RP) Panel. The BioFire RP Panel is a multiplex nested RTPCR test capable of detecting up to 20 respiratory pathogens found in respiratory samples obtained from patients exhibiting respiratory disease symptoms. PER incorporates real-time polymerase chain reaction metrics from the BioFire RP Panel RV/EV assays as model inputs. PER was tuned and tested using data from 1,619 samples spanning two years, from six institutions in the United States and Europe. The samples were confirmed for EV-D68 by gold-standard molecular methods at the six institutions. The final PER algorithm demonstrated an overall sensitivity and specificity of 87.1% and 86.1%, respectively, among the gold-standard dataset. PER works in conjunction with BioFire^®^ (Trend), an epidemiology network that connects participating BioFire^®^ Syndromic Trends Systems to a centralized database, to predict rates of EV-D68 on a population level. PER has been used as a real time monitoring tool to alert participating sites of predicted EV-D68 circulation and was in effect during the previous 2018 outbreak [1].

Monthly and annual time series of confirmed AFM cases is obtained from the CDC website (https://www.cdc.gov/acute-flaccid-myelitis/cases-in-us.html. Accessed February 11, 2020). The monthly time series is extracted from the website using WebPlotDigitizer [2].

### S1.2 Demographic data

The monthly numbers of births and deaths in each state between 2014–2017 are obtained from the Centers for Disease Control and Prevention (CDC) Wonder database [3; 4]. The population sizes in each state and in each city (only including the cities from which samples were collected) are obtained from the Census Bureau website [5]. Birth and death rates are estimated by dividing the monthly numbers of births and deaths by the total population size in each state and interpolating with cubic splines. For our epidemiological model, we adjust the population size to account for the cities from which samples were collected by using the sum of population sizes of those cities, instead of the population size of each state.

### S1.3 Variance stabilization of a time series

Before we measure cross-correlations or perform wavelet analyses of time series, we transform the time series to stabilize their variances. Integers (e.g., number of reported AFM cases) are log-transformed after adding 1: log(*x*+1) [6]. Proportions (e.g., proportion of PER-predicted EV-D68 cases) are arcsine-square-root transformed: arcsin 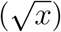 [7].

### S1.4 Wavelet analysis

We perform Morlet wavelet analyses of the arcsine-square-root transformed proportion of PER-predicted EV-D68 cases to characterize the periodicity of the observed EV-D68 cycles. Confidence intervals are calculated from 100 bootstrap samples. The analyses are performed using the “WaveletComp” `R` package.

### S1.5 Center of gravity

The center of gravity is calculated by taking the average of weeks in each even year (2014, 2016, and 2018), weighted by the proportion of PER-predicted EV-D68 cases, and then taking the mean of these averages [8]. We only use epidemics that occur during the even years to characterize the patterns of major epidemics. Center of gravity is calculated using circular statistics with the “circular” `R` package [9].

### S1.6 Epidemiological model

We use a homogeneous, stochastic Susceptible-Infected-Recovered (SIR) model with realistic birth and death rates to describe the spread of EV-D68 in each state [10] and fit the model to weekly proportion of PER-predicted EV-D68 cases. The model is an Euler approximation of a continuous-time SIR model [11]. For simplicity, we discretize both the simulation and the observation time steps at a scale of 1 week (Δ*t* = 1 week):

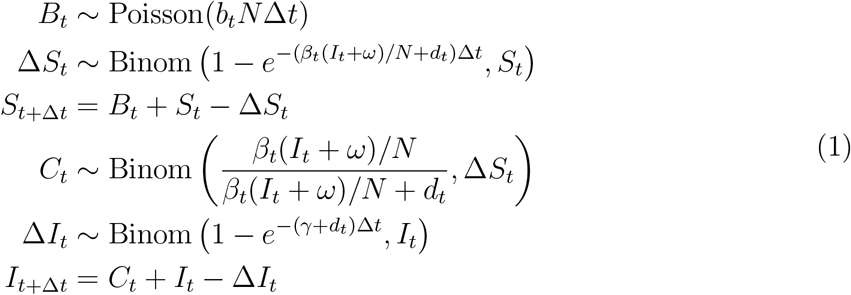

where *b*_*t*_ and *d*_*t*_ are the estimated time-varying birth and death rates, *β*_*t*_ is the time-varying transmission rate, *γ* is the recovery rate, and *ω* is the constant external forcing term. For computational efficiency, we do not keep track of the recovered population *R* and assume that the population *N* stays constant. This does not mean that we are letting the birth and death rates equal; the birth and death rates vary over time to reflect the observed number of birth and death. We are simply keeping the *N* in the denominator of the force of infection (*βI/N*) and in the total expected birth *b*_*t*_*N* constant. Given that our time series is very short, assuming a constant *N* has negligible effects on the epidemiological dynamics as well as parameter estimates.

We model transmission rate *β*_*t*_ using cyclic B-splines with 5 knots, at intervals of 1/4 years starting at 2014:

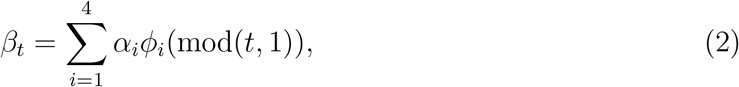

where *α*_*i*_ represent spline coefficients and *ϕ*_*i*_ represent spline bases. The mean transmission rate is given by the mean of the spline coefficients. The basic reproduction number ℛ_0_ is calculated using the mean transmission rate while accounting for the discretization step [11]:

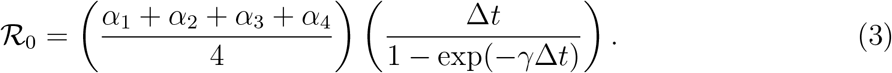

We initially used a sinusoidal function to model the transmission rate but found that it was not able to capture the steepness of epidemic curves.

In order to model the probability of detecting a positive case in a sample, we first assume that there is an average, baseline rate *ρ*_0_ at which the total population gets tested. Then, the average total number of individuals that get tested between time *t* and *t* + Δ*t* is given by

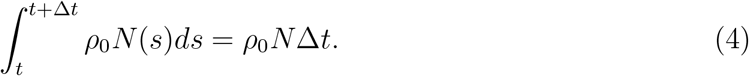

On the other hand, we assume that newly infected individuals get tested at a rate *ρ*_1_. Then, the probability of detecting EV-D68 in a sample at time *t* is given by

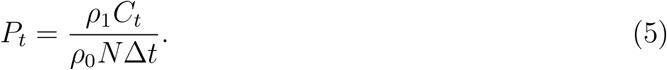

We reparameterize this model by writing *λ* = *ρ*_1_*/ρ*_0_, which represents the relative rate at which individuals infected with EV-D68 get tested compared to the rest of the population. Although it is possible to explicitly model individuals that have already been tested and those that have not yet been tested, our method provides a simple, but reasonable, approximation when the prevalence is low and the generation time is short.

Finally, we model the observed number of EV-D68 cases *C*_obs,t_ using a beta-binomial model:

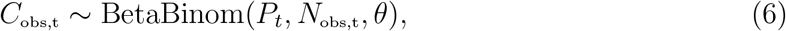

where *N*_obs,t_ represent the observed number of individuals tested positive for RV/EV at time *t*, and *θ* represents the Beta-binomial over-dispersion parameter. This observation model assumes that there is a constant rate at which individuals, on average, get tested for RV/EV; in reality, we expect the testing rate to fluctuate over time depending on the seasonalities of RV/EV serotypes. We are unable to model this fluctuation explicitly because we have incomplete information about the observed RV/EV serotypes within samples. We implicitly account for this fluctuation in the beta-binomial likelihood by incorporating the observed number of RV/EV samples. We implement the model using the R package `pomp` [12].

### S1.7 Inferential framework

Following [10], we assume *γ* = 1*/*7 days^*−*1^. Then, we try to find the maximum likelihood estimates (MLEs) of the remaining model parameters: initial proportion of infected individuals *I*(0) at the beginning of 2014, initial proportion of susceptible individuals *S*(0) at the beginning of 2014, four spline coefficients *α*_1_, *α*_2_, *α*_3_, *α*_4_, external forcing term *ω*, relative rate at which individuals infected with EV-D68 get tested *λ*, and the beta-binomial dispersion parameter *θ*. In order to find the MLEs, we first construct 50 starting parameter sets for the model that assumes a sinusoidal transmission rate, i.e., *β*_*t*_ = *β*_*m*_(1 + *β*_*s*_ cos(2*π*(*t* + *A*))), using Latin Hypercube Sampling (LHS). For each LHS sample, we perform trajectory matching. Starting from the parameter set estimated from the trajectory matching, we fit the stochastic model using the iterated, perturbed Bayes map (IF2) algorithm [13]. We use 300 MIF iterations with 5000 particles during this step.

For each stochastic fit using a sinusoidal transmission rate, we translate the sinusoidal function parameters (*β*_*m*_, *β*_*s*_ and *A*) into spline coefficients by performing a linear regression using the estimated time-varying transmission rates as a response variable and spline bases as covariates. We fit the stochastic model with spline-based transmission rates using the IF2 algorithm, starting from the parameters we estimated from the previous step. We use 600 MIF iterations with 5000 particles during this step. For each fit, we calculate the log-likelihood by taking logarithm of the mean of the 10 replicates of the particle-filter likelihoods, each using 10000 particles. The resulting parameter set with the highest log-likelihood is the MLEs of the model parameters. We expect this two-step approach to converge better than a one-step approach that tries to estimate spline coefficients directly from LHS samples because the latter approach could give unrealistic shapes of the transmission rates as starting points.

To calculate the 95% confidence intervals associated with each parameter, we use importance sampling [14]. We first transform the MLEs of the model parameters to unconstrained scales: parameters confined to positive values are log-transformed, and those confined between 0 and 1 are logit-transformed. We draw 5000 random parameter sets from a multivariate normal distribution, using the transformed MLEs as the mean, and calculate the particle-filter likelihood at each point using 10000 particles. The weight of each parameter set is calculated by dividing the particle-filter likelihood by the sampling probability density of the parameter set. The weighted parameter samples give an approximate posterior distribution that assumes flat priors on all parameters on unconstrained scales. We use the scaled variance-covariance matrix of the parameter sets that were proposed during IF2 steps, which are less than 4 log-likelihood units away from the log-likelihood of the MLEs, as the variance-covariance matrix of the multivariate normal proposal distribution. The scaling factors for the variance-covariance matrix are determined through trial-and-error so that the importance samples can explore a sufficiently wide range of parameters; the scaling factors vary between 10 and 100 across different regions. Confidence intervals are calculated by taking the weighted quantiles of the approximate posterior distribution.

To validate importance sampling confidence intervals, we compare them with the approximate profile confidence intervals. Approximate log-likelihood profiles are calculated by dividing the marginal importance samples into 50 equi-spaced bins on unconstrained scales and taking the maximum log-likelihood in each bin. We fit a Locally Estimated Scatterplot Smoothing (LOESS) regression to maximum log-likelihoods across 50 bins and calculate the confidence intervals for each parameter from the estimated log-likelihood profile curves from the LOESS regression. Both methods give consistent results (Fig. S16–S21).

### S1.8 Bifurcation diagram

Long-term dynamics of childhood diseases and their dynamical transitions can be predicted from bifurcations of a deterministic model [15; 16]. Here, we use a bifurcation diagram to study long-term stability of the observed biennial cycles. For each combination of relative amplitude and *ℛ*_0_, we run the deterministic model for 200 years (in order to remove any transient dynamics) assuming equal birth and death rates (using birth rates from New York, 2014) to keep the population at demographic equilibrium. Then, we calculate the mean periodicity from the wavelet analysis of the final 100 years of logged incidence. The estimated *ℛ*_0_ for each state is adjusted by multiplying the relative average birth rate, compared to the average birth rate in New York.

## S2 Supplementary Text

### S2.1 Geographical differences in EV-D68 and AFM surveillance

The biggest numbers of AFM cases are reported from California (89 out of 606 total cases), where the observed peak month matches the observed peak month at the national level [17], and Texas (63 out of 606 cases). PER-predicted EV-D68 reports from the BioFire^*®*^ Trend are concentrated in Northeast and Midwest regions (6534 out of 10395 total cases), particularly in New York (2478 out of 10395 total cases), but are sparse in California and Texas (446 out of 10395 total cases). These differences are likely to play important roles in shaping the differences in seasonality of the PER-predicted EV-D68 dynamics and the observed AFM patterns at the national level.

### S3 Supplementary Figures

**Figure S1:**
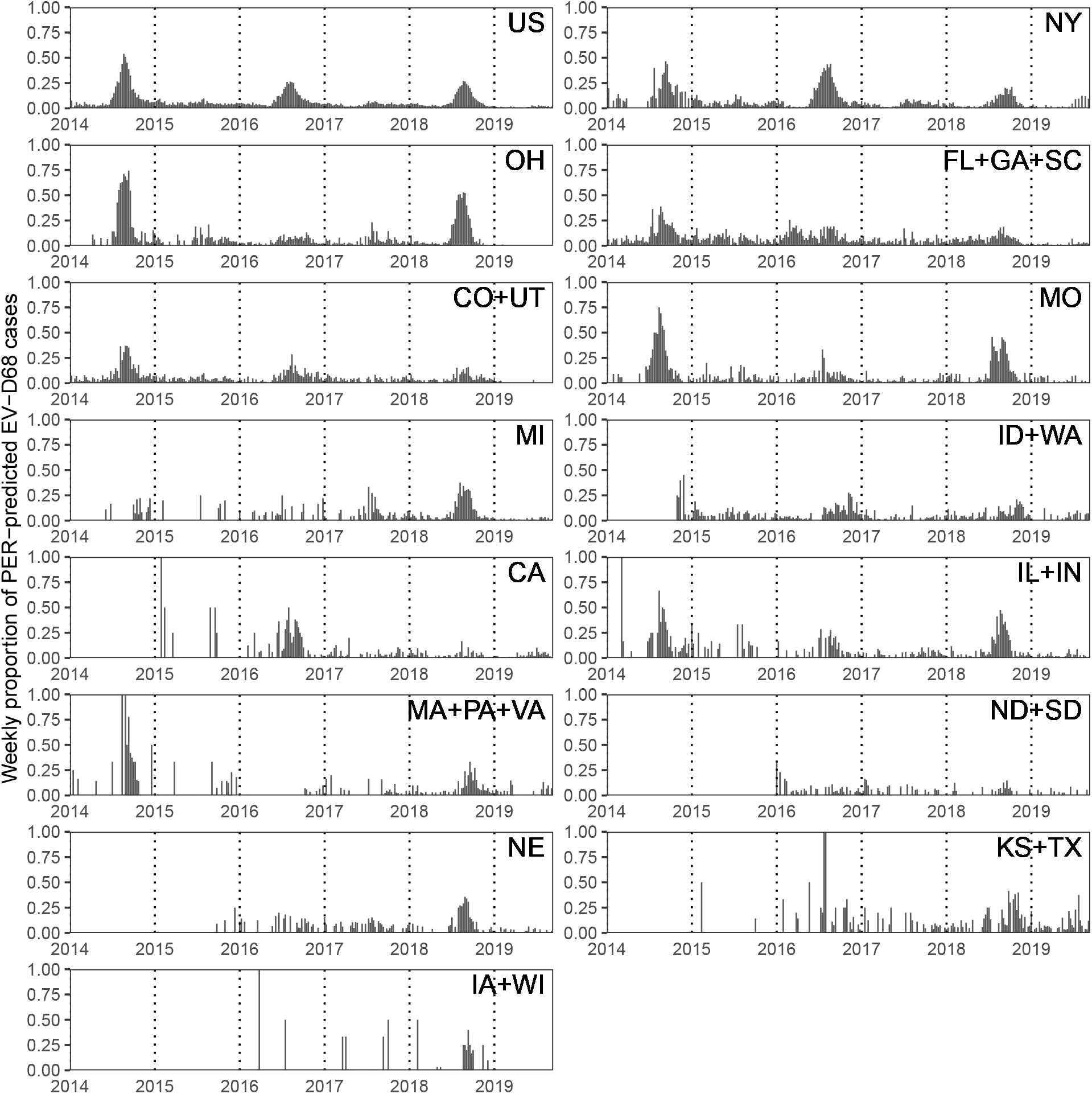
Weekly proportion of PER-predicted EV-D68 cases in the US and in 14 regions. Regions are ranked by the total number of RV/EV samples.

**Figure S2:**
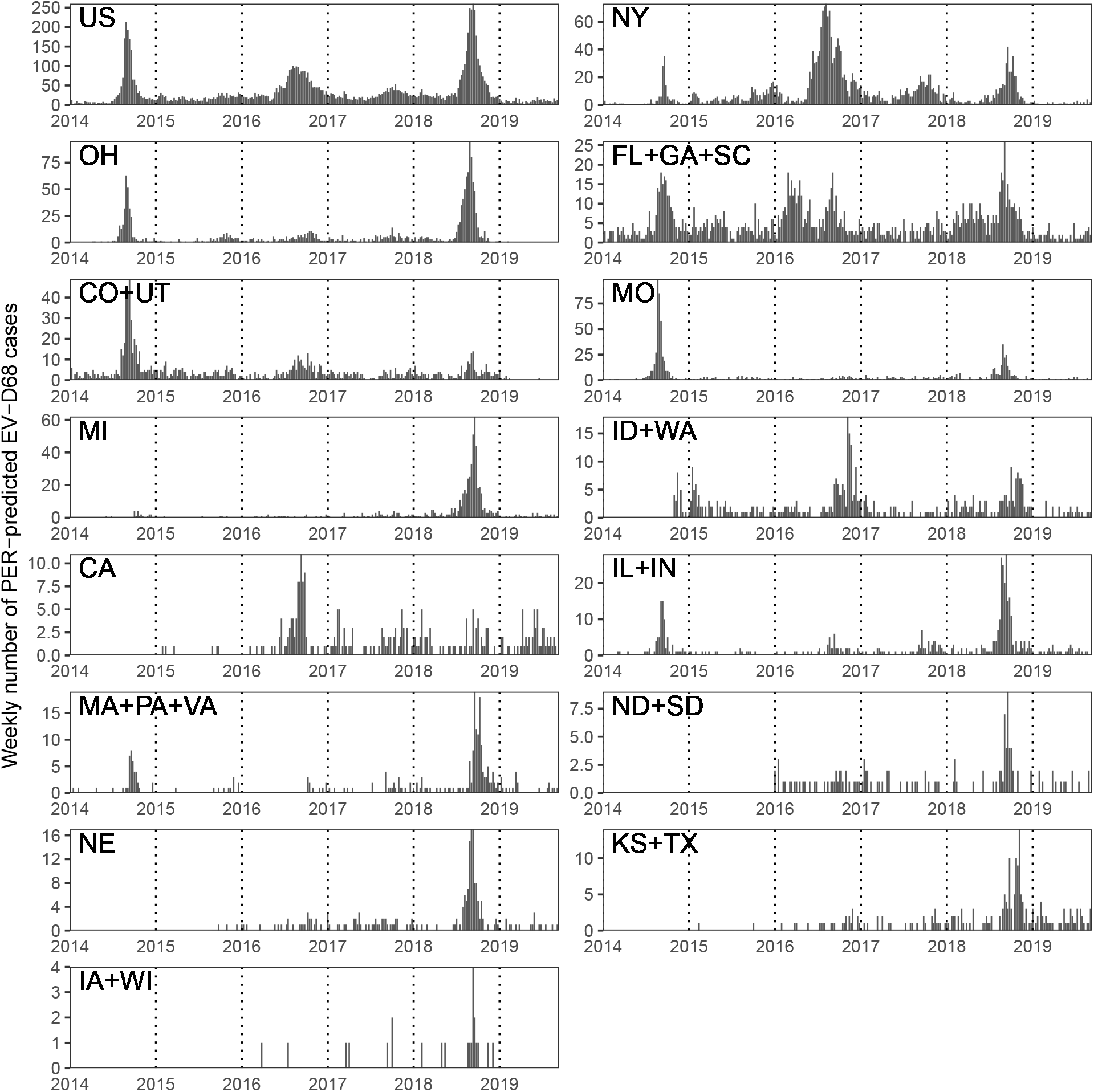
Weekly number of PER-predicted EV-D68 cases in the US and 14 regions.

**Figure S3:**
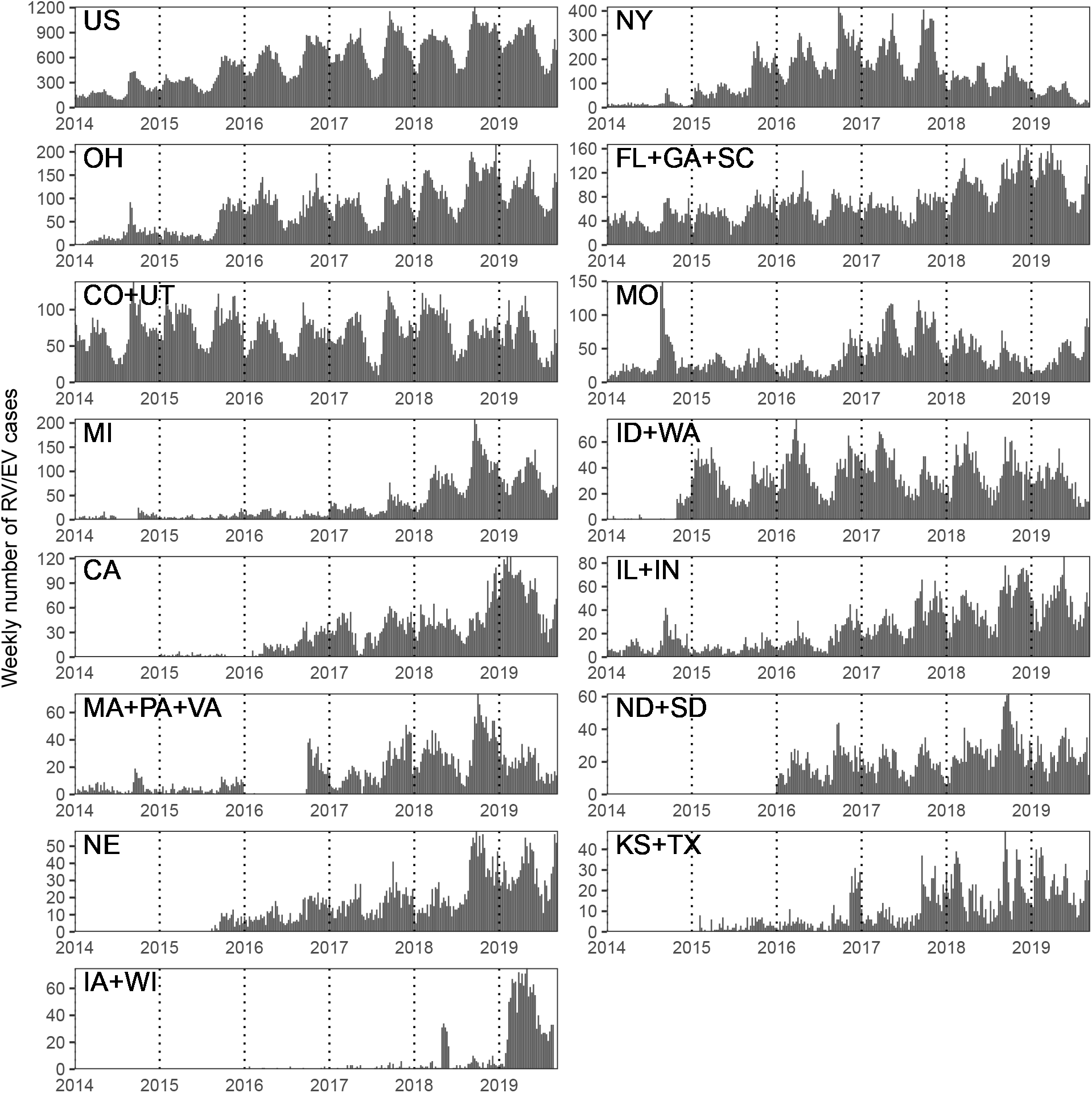
Weekly number of RV/EV samples in the US and 14 regions.

**Figure S4:**
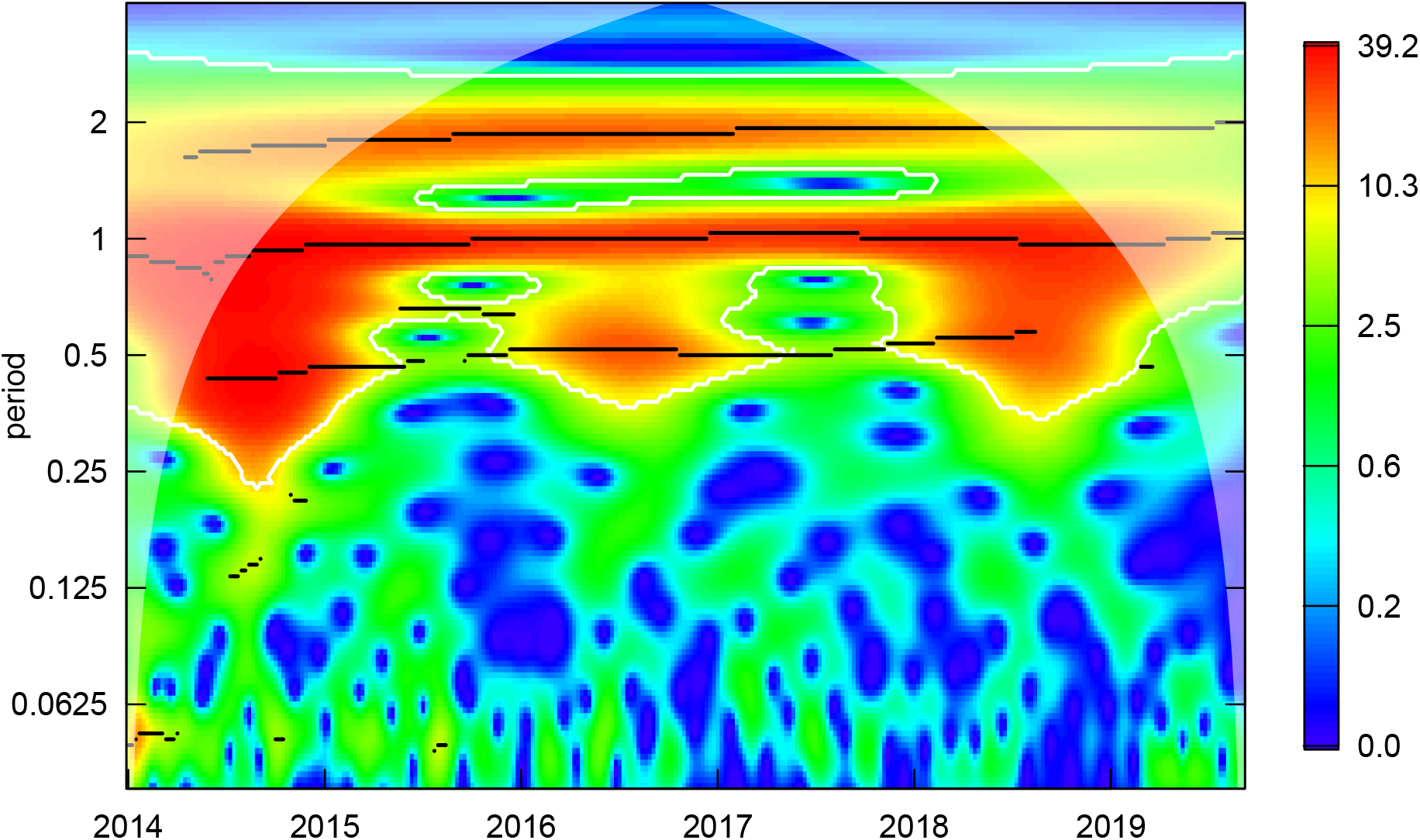
Result of the wavelet analysis of the total variance stabilized EV-D68 time series in the US. The white lines represent the confidence intervals calculated from bootstrap samples. The black lines represent the wavelet ridge. The white shades represent the cone of influence.

**Figure S5:**
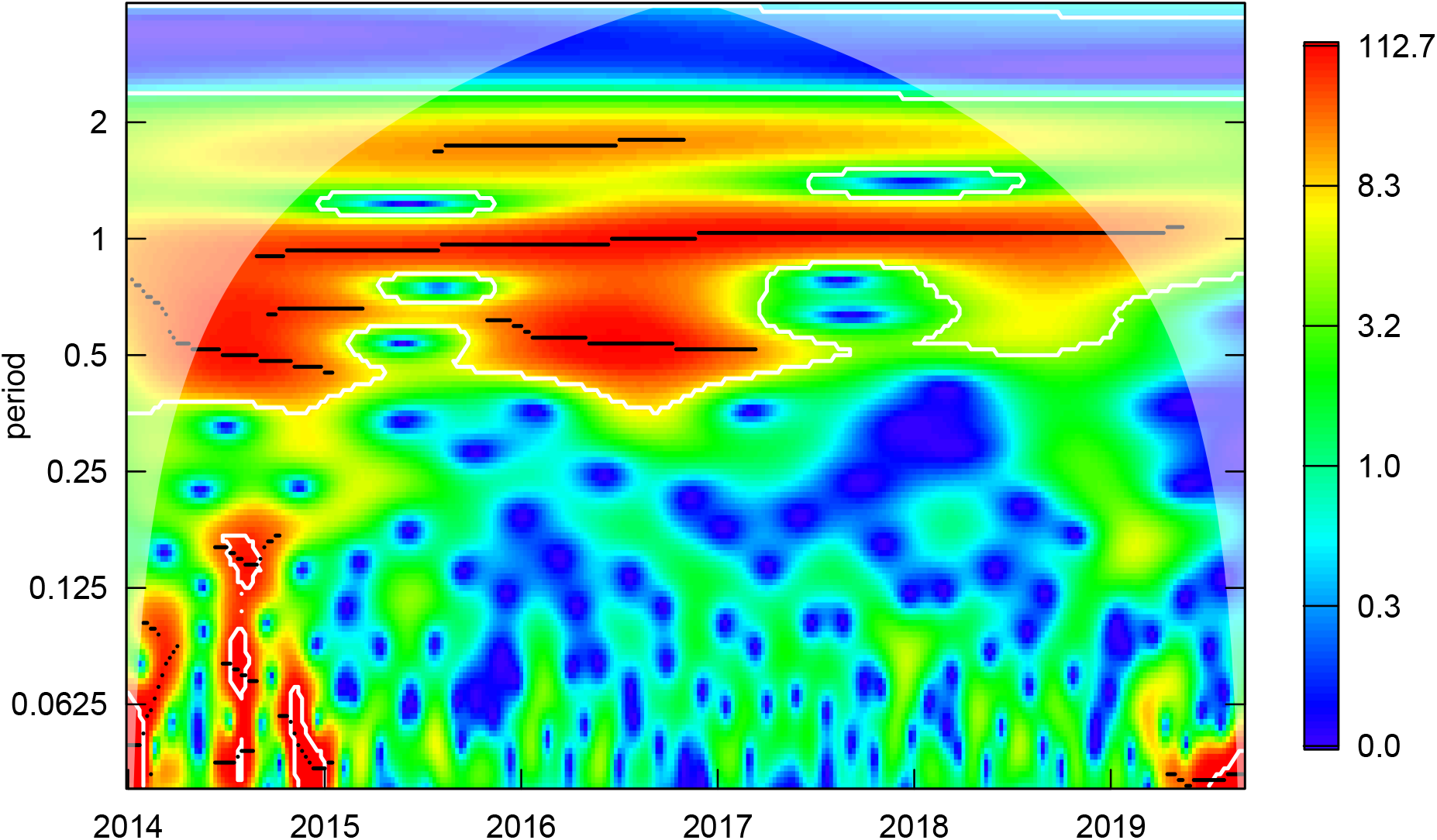
Result of the wavelet analysis of the variance stabilized EV-D68 time series in New York. The white lines represent the confidence intervals calculated from bootstrap samples. The black lines represent the wavelet ridge. The white shades represent the cone of influence.

**Figure S6:**
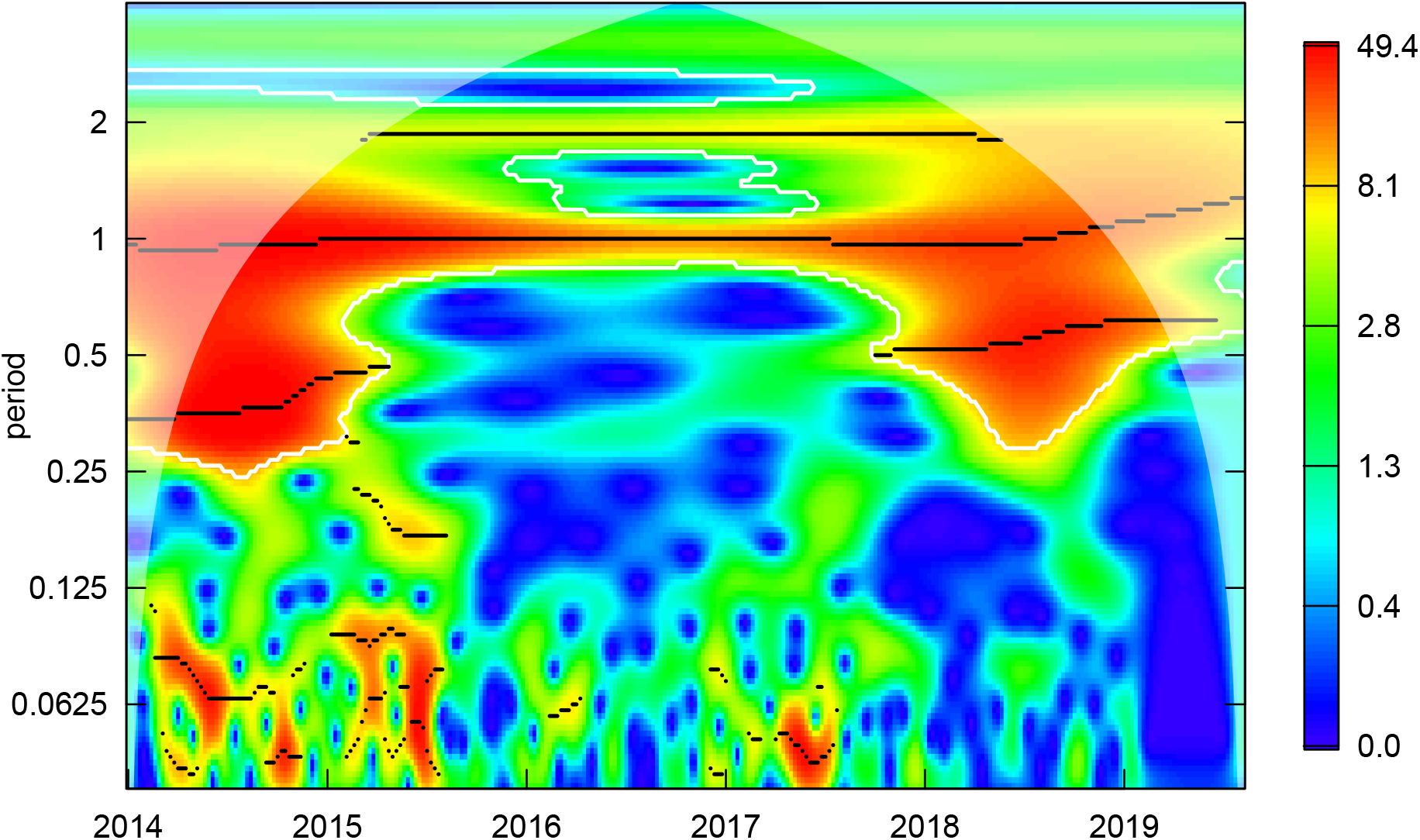
Result of the wavelet analysis of the variance stabilized EV-D68 time series in Ohio. The white lines represent the confidence intervals calculated from bootstrap samples. The black lines represent the wavelet ridge. The white shades represent the cone of influence.

**Figure S7:**
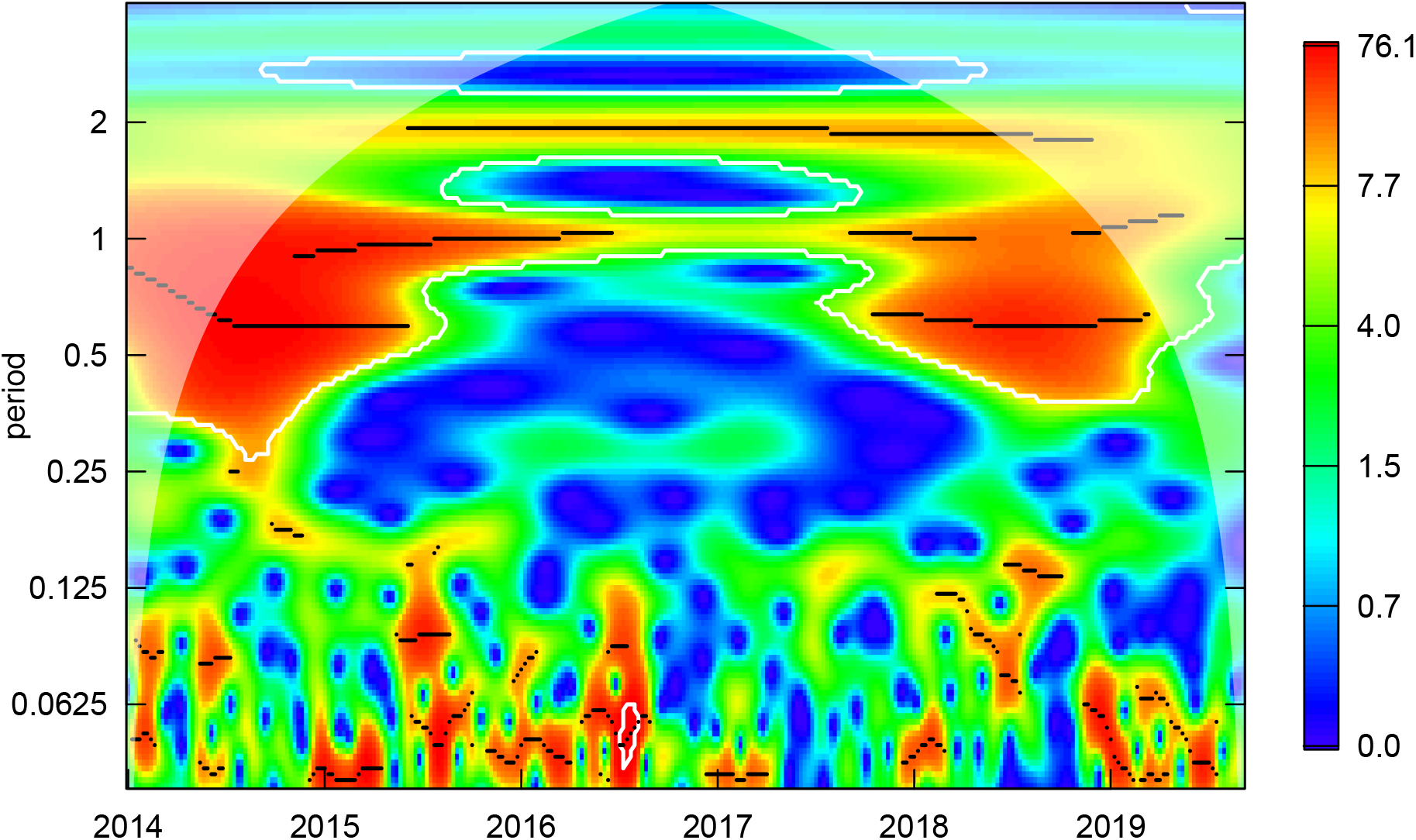
Result of the wavelet analysis of the variance stabilized EV-D68 time series in Missouri. The white lines represent the confidence intervals calculated from bootstrap samples. The black lines represent the wavelet ridge. The white shades represent the cone of influence.

**Figure S8:**
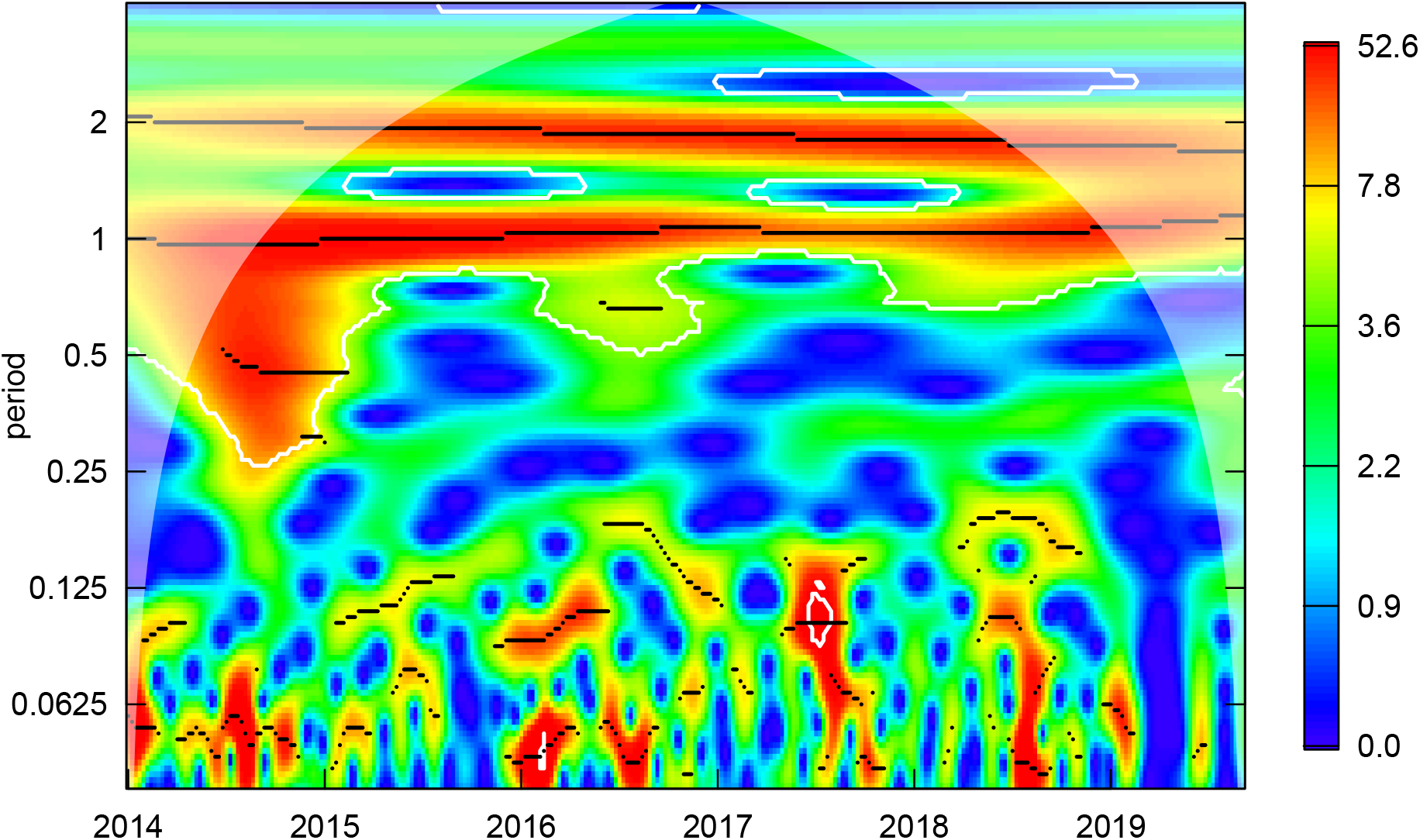
Result of the wavelet analysis of the variance stabilized EV-D68 time series in the Utah and Colorado and region. The white lines represent the confidence intervals calculated from bootstrap samples. The black lines represent the wavelet ridge. The white shades represent the cone of influence.

**Figure S9:**
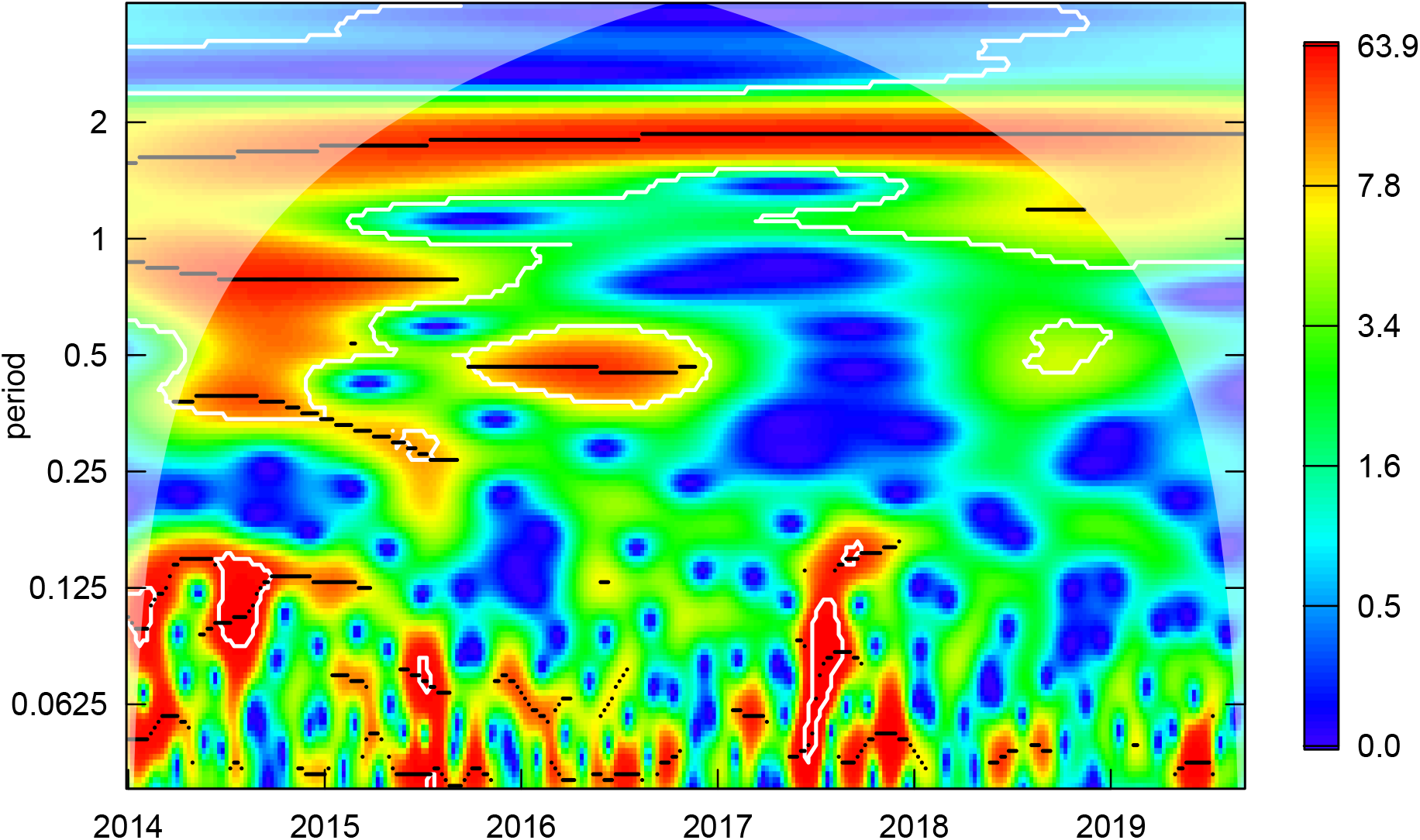
Result of the wavelet analysis of the variance stabilized EV-D68 time series in the Florida, Georgia, and South Carolina region. The white lines represent the confidence intervals calculated from bootstrap samples. The black lines represent the wavelet ridge. The white shades represent the cone of influence.

**Figure S10:**
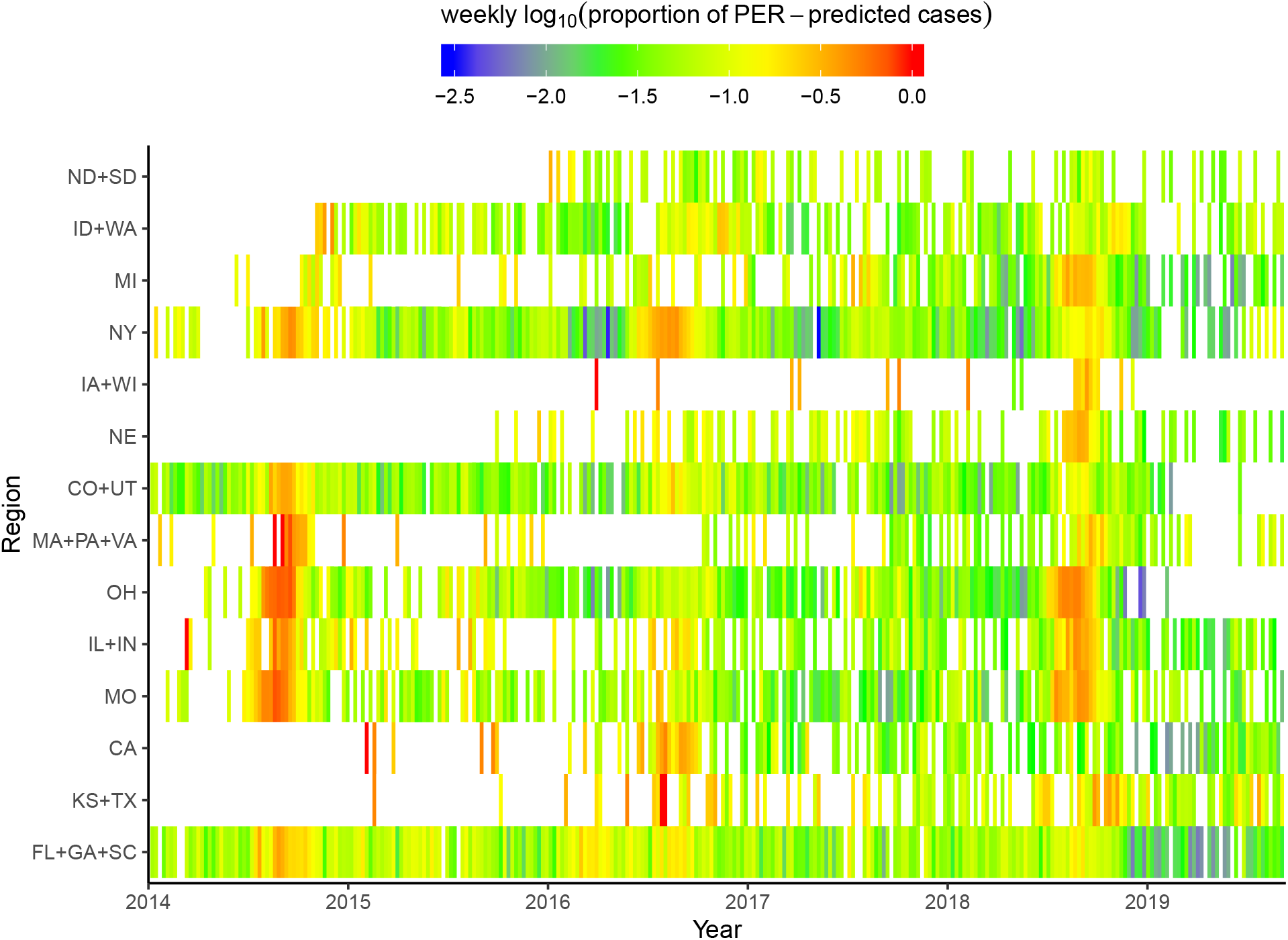
A comparison of weekly proportion of PER-predicted EV-D68 cases across 14 regions. Thick orange bands in 2014 and 2018 suggest that the outbreaks are synchronized in many states. A lack of orange band in 2016 suggests that the outbreaks are not synchronized. Regions are ranked by the mean latitude of the capital city of each states.

**Figure S11:**
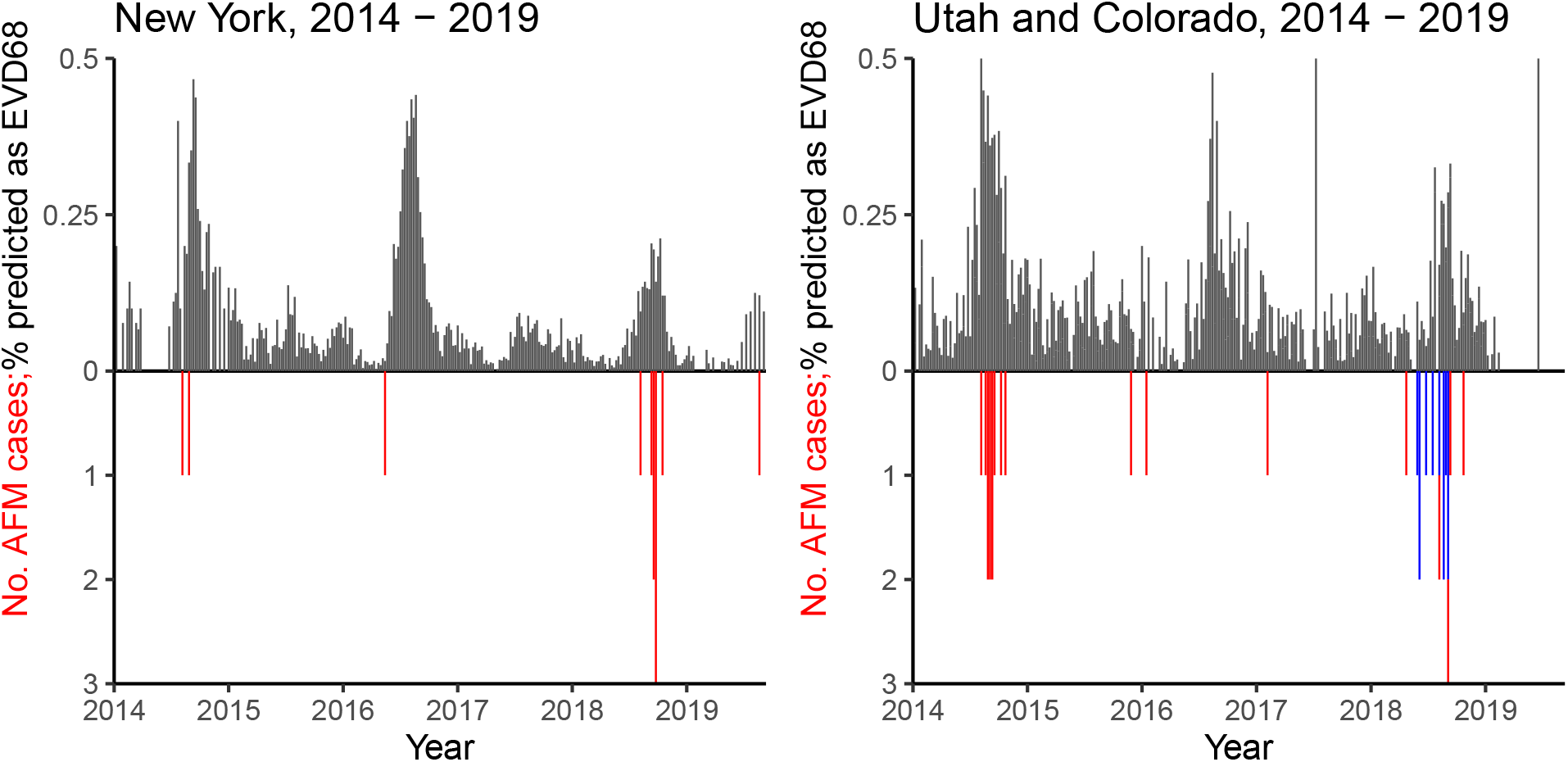
A fine scale comparison of weekly proportion of PER-predicted EV-D68 cases and weekly number of AFM cases in New York and Colorado. The black bars represent the weekly proportion of PER-predicted EV-D68 cases. The red bars represent the weekly number of AFM cases, except for those that are known to be associated with EV-A71. The blue bars represent weekly number of AFM cases that are known to be associated with EV-A71. AFM reports from New York and Colorado are obtained from the New York State Department of Health and Colorado Department of Public Health and Environment, respectively. While the AFM cases are from Colorado in the right panel, we present PER-predicted proportion of EV-D68 cases from both Utah and Colorado due to confidentiality requirements.

**Figure S12:**
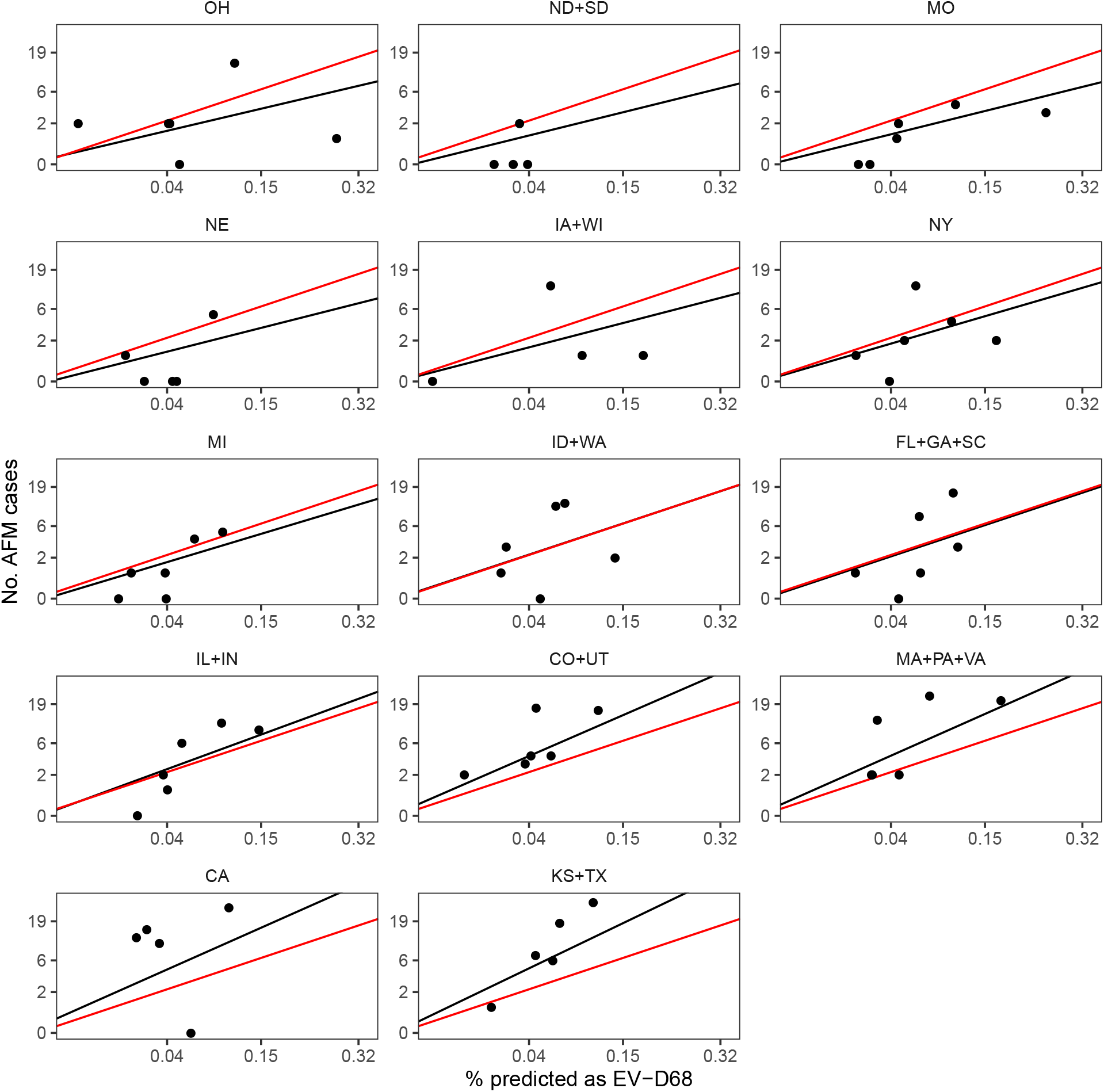
A comparison of annual proportion of PER-predicted EV-D68 cases and annual number of AFM cases across 14 regions with lines estimated from a linear mixed model with random slopes and intercepts. Black lines represent the individual-level prediction using random effect estimates (conditional modes). Red lines represent the population-level prediction using fixed effect estimates. Regions are ranked by the estimated random slopes.

**Figure S13:**
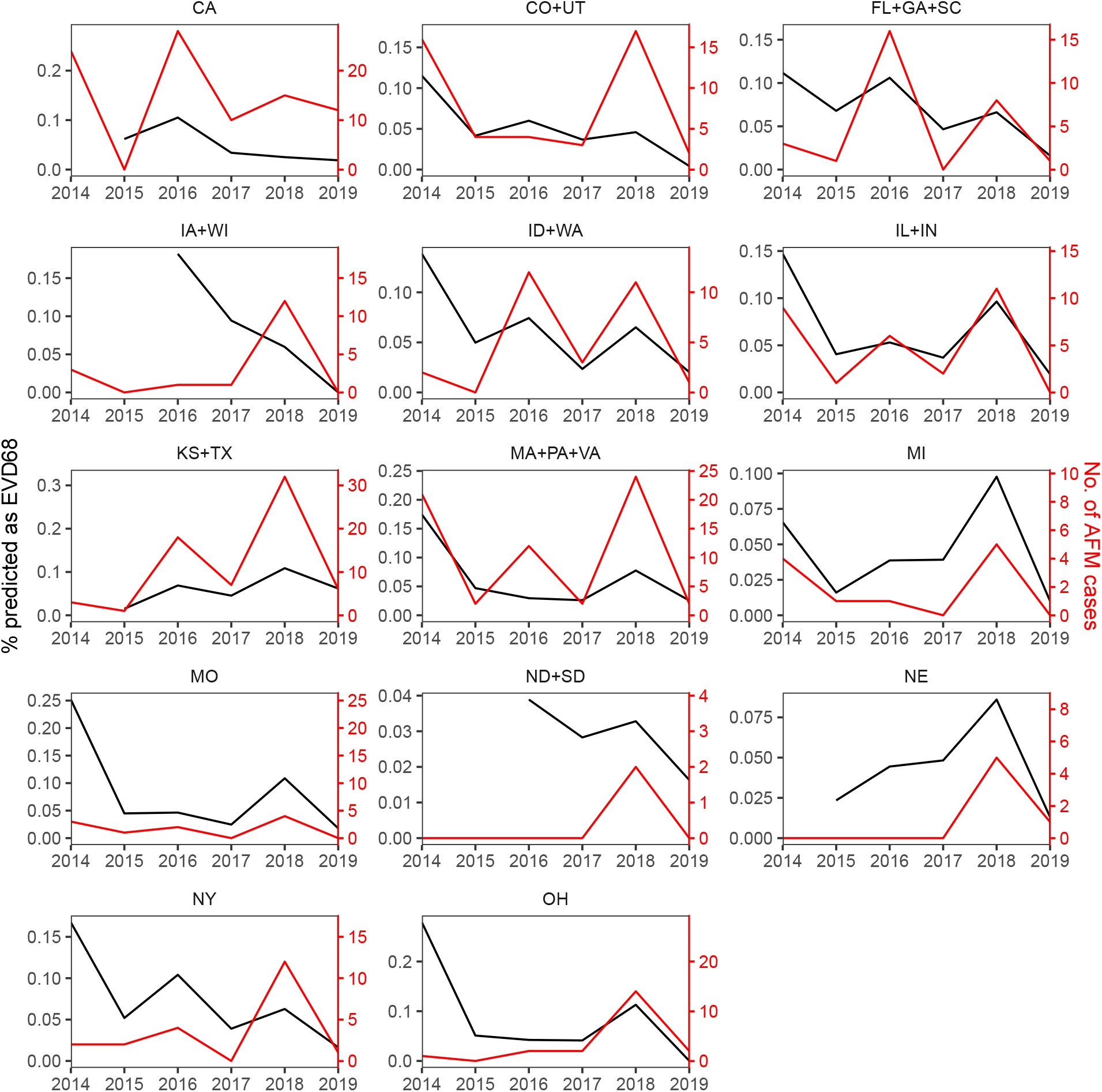
A comparison of annual proportion of PER-predicted EV-D68 cases and annual number of AFM cases across 14 regions. Regions are ranked alphabetically.

**Figure S14:**
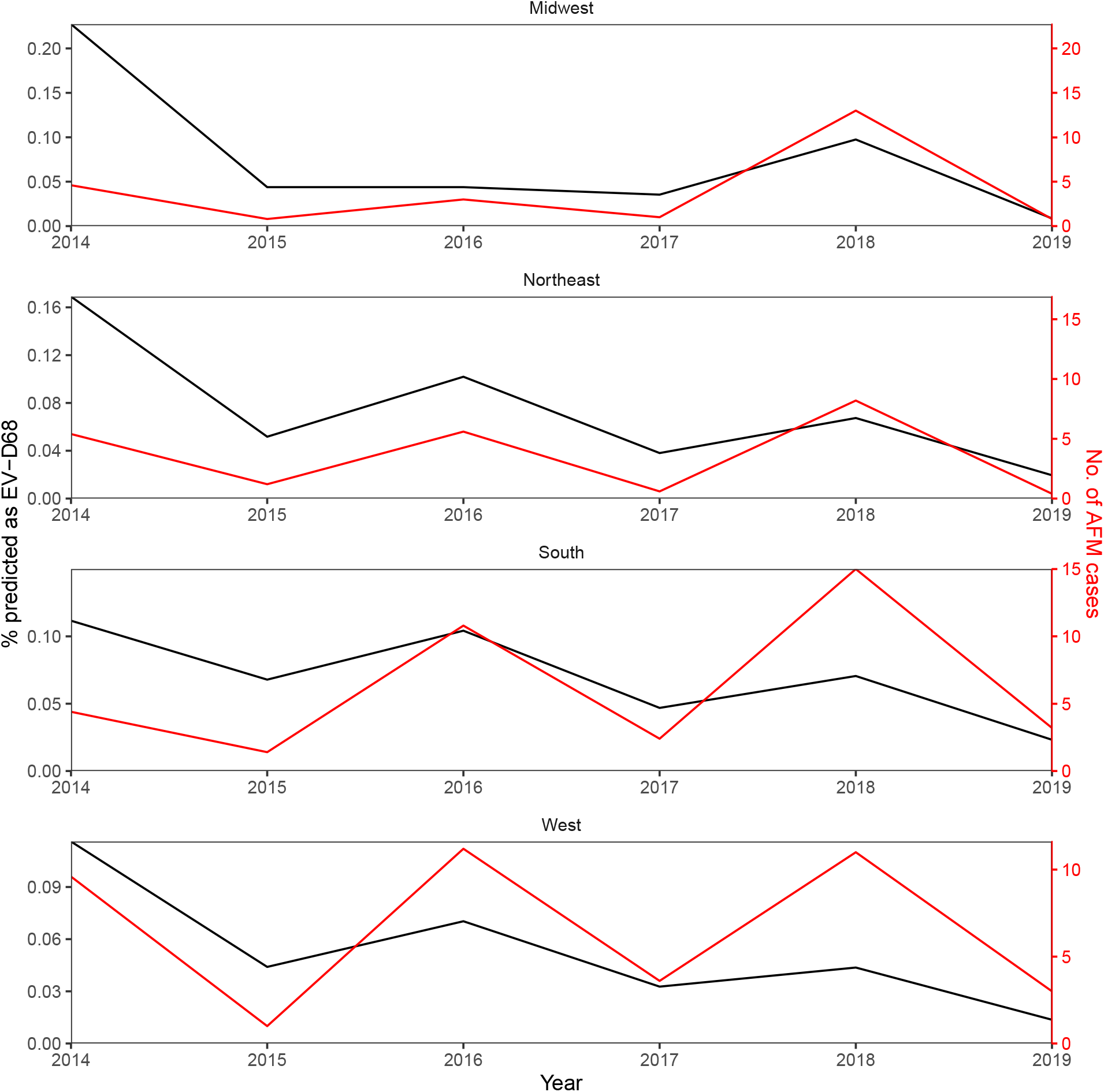
A comparison of annual proportion of PER-predicted EV-D68 cases and annual number of AFM cases across 4 census regions.

**Figure S15:**
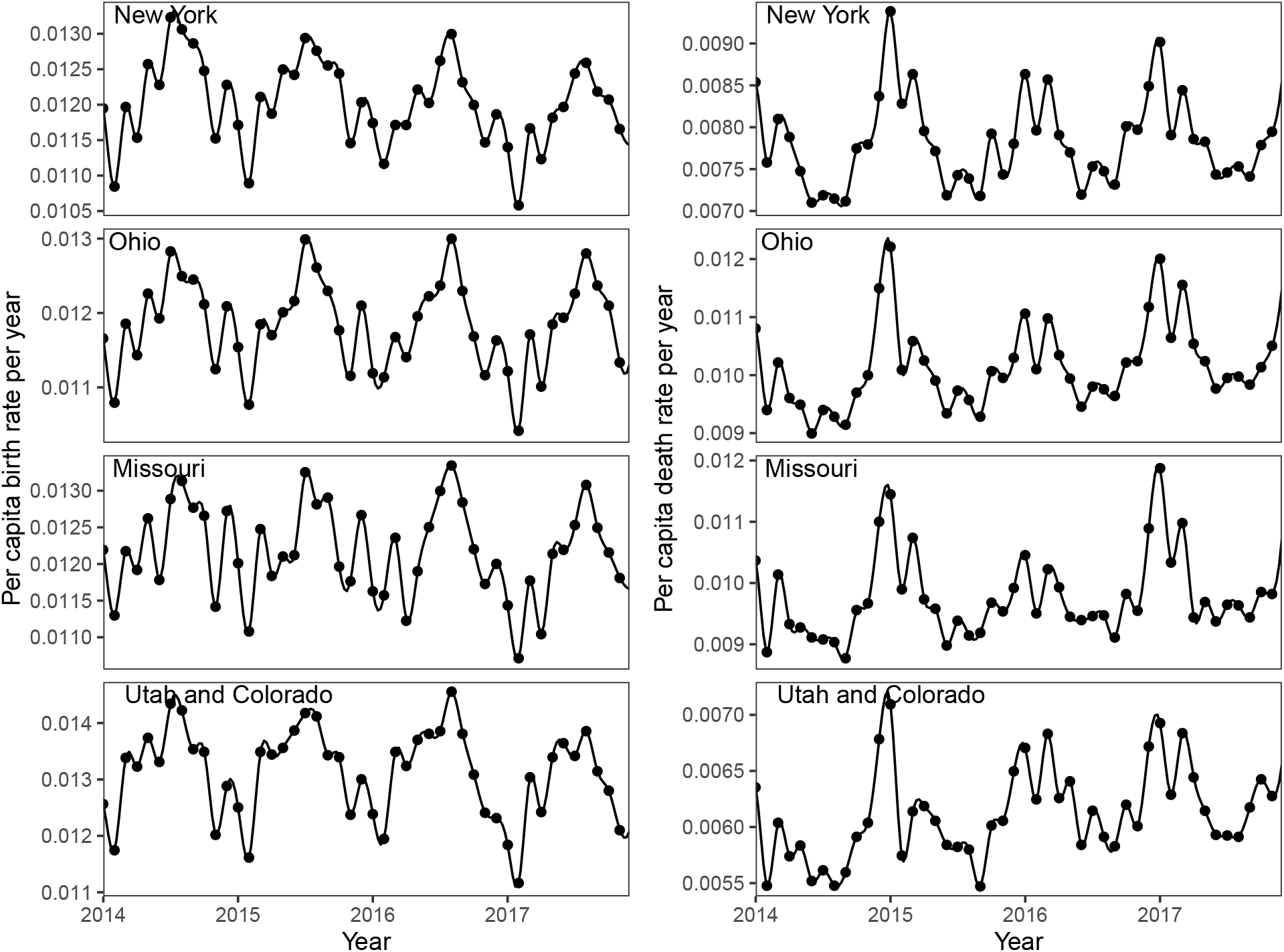
Per capita birth and death rates per year. The black points represent the observed birth and death rates. The black lines represent the estimated birth and death rates using cubic splines.

**Figure S16:**
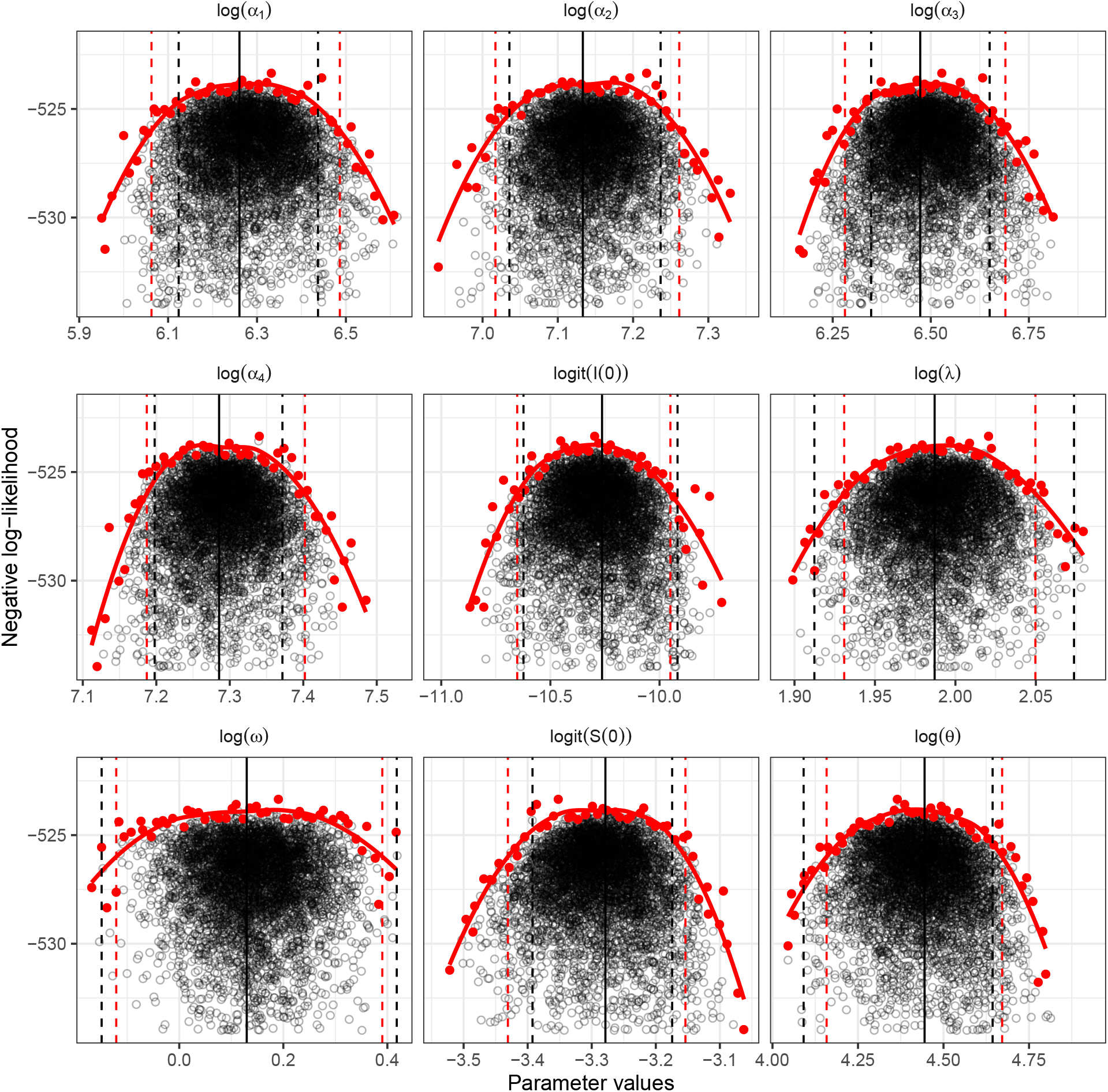
Parameter estimates and confidence intervals for New York. The black, solid, vertical lines represent the MLEs. The black, dashed, vertical lines represent the 95% confidence intervals calculated by importance sampling. The red, dashed, vertical lines represent the 95% confidence intervals calculated by approximate profile likelihood. Black points show importance samples. Only parameter sets within 10 log-likelihood units from the maximum likelihood are shown.

**Figure S17:**
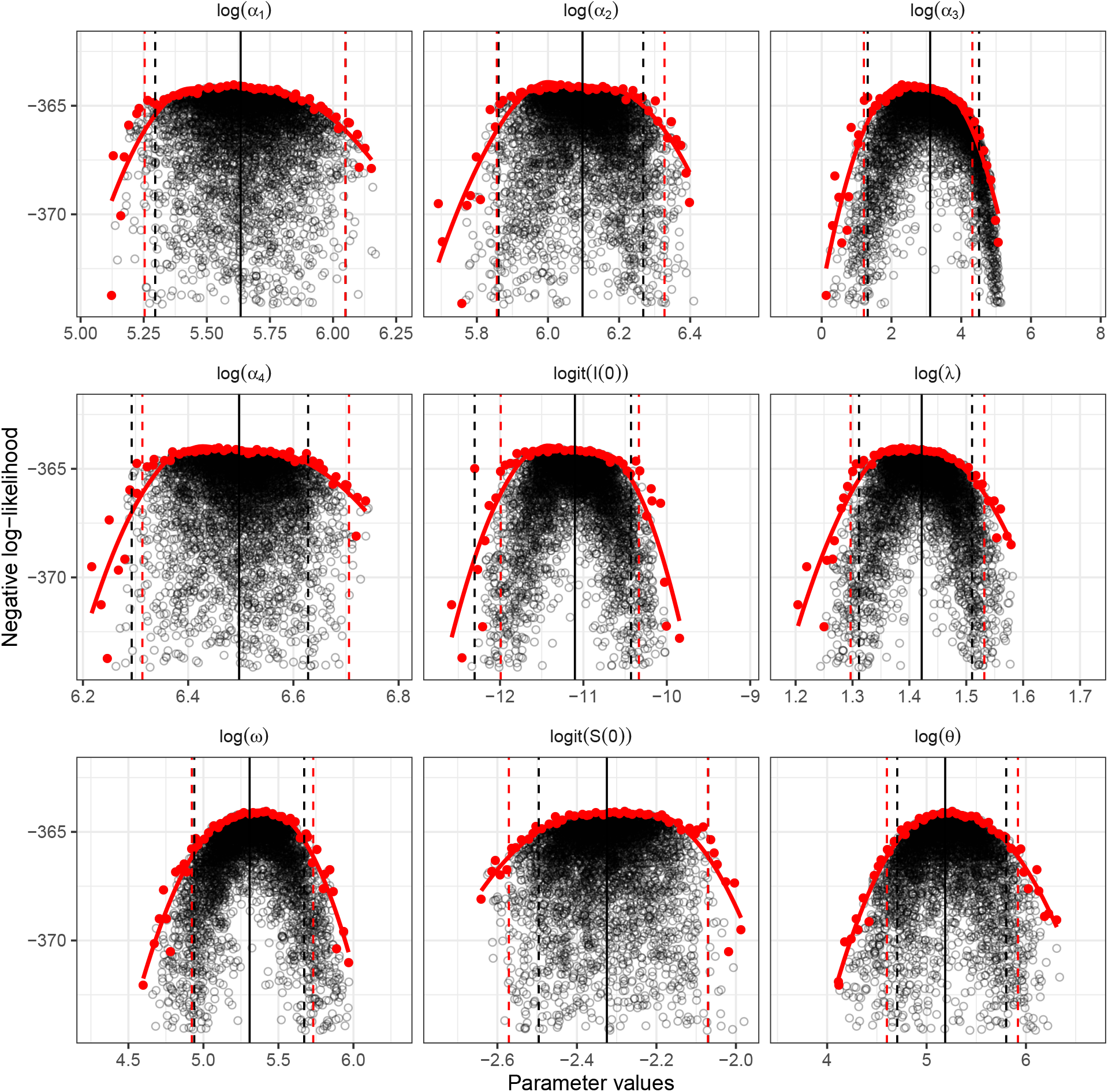
Parameter estimates and confidence intervals for Ohio. The black, solid, vertical lines represent the MLEs. The black, dashed, vertical lines represent the 95% confidence intervals calculated by importance sampling. The red, dashed, vertical lines represent the 95% confidence intervals calculated by approximate profile likelihood. Black points show importance samples. Only parameter sets within 10 log-likelihood units from the maximum likelihood are shown.

**Figure S18:**
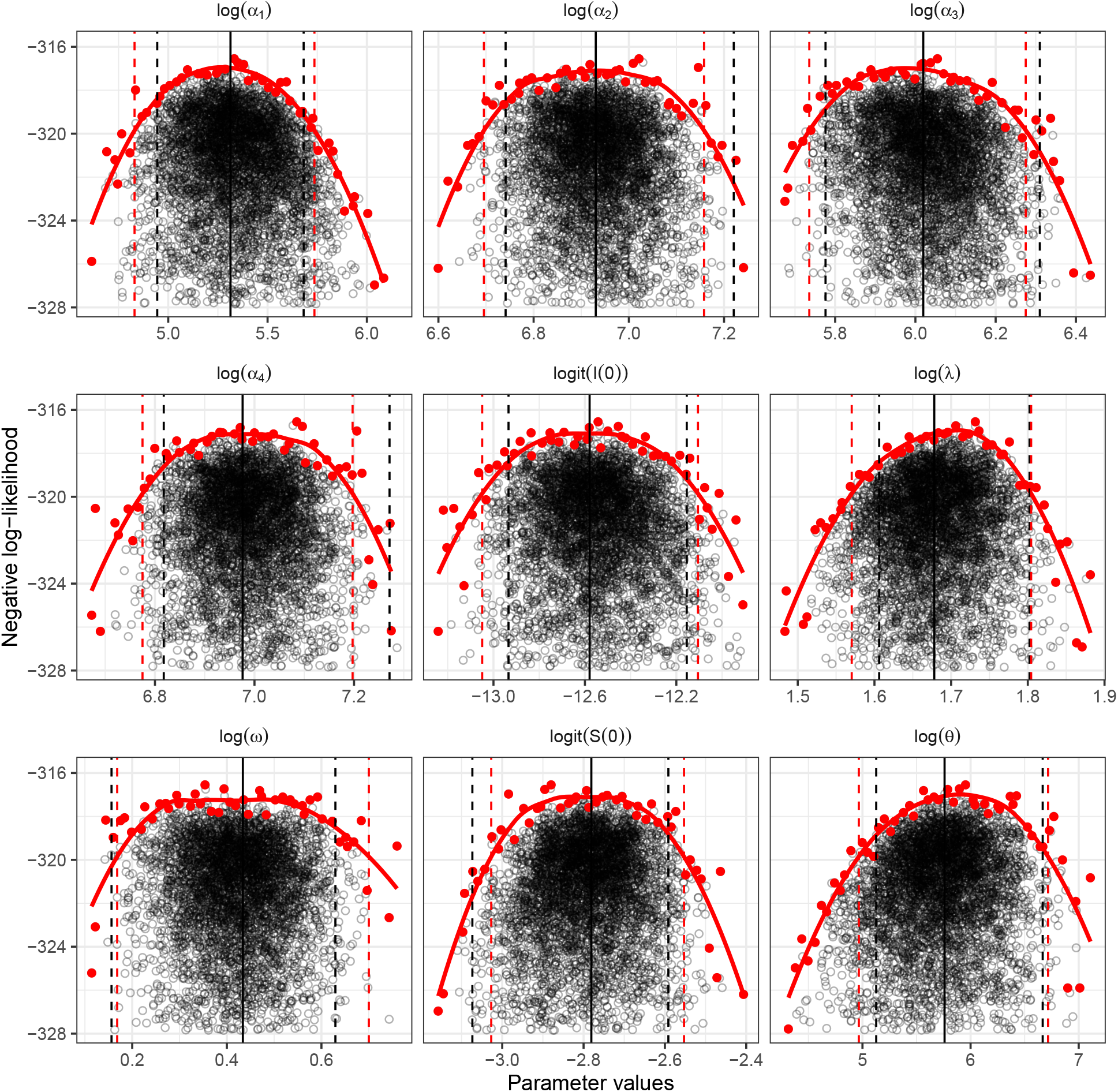
Parameter estimates and confidence intervals for Missouri. The black, solid, vertical lines represent the MLEs. The black, dashed, vertical lines represent the 95% confidence intervals calculated by importance sampling. The red, dashed, vertical lines represent the 95% confidence intervals calculated by approximate profile likelihood. Black points show importance samples. Only parameter sets within 10 log-likelihood units from the maximum likelihood are shown.

**Figure S19:**
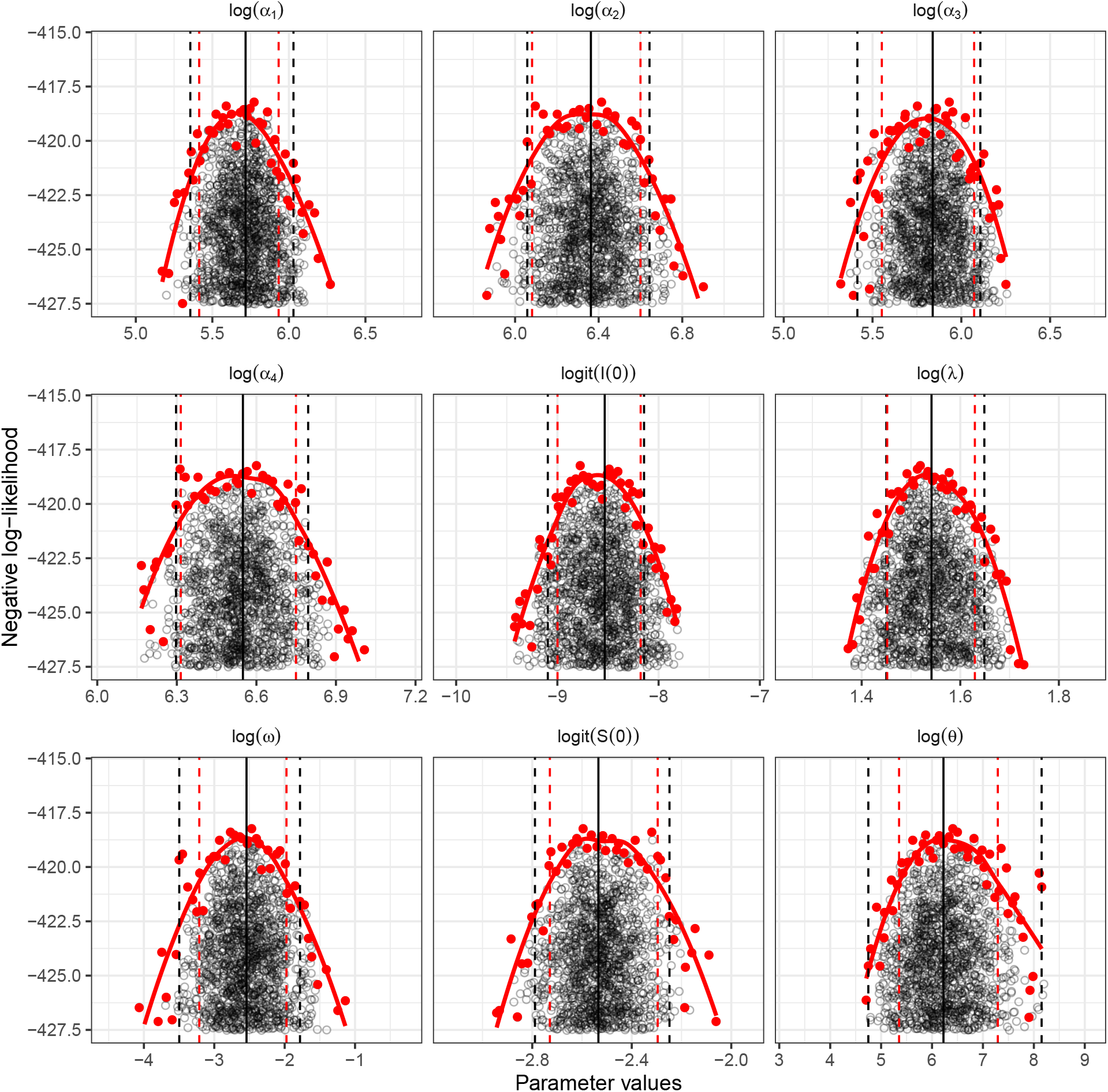
Parameter estimates and confidence intervals for the Utah and Colorado region. The black, solid, vertical lines represent the MLEs. The black, dashed, vertical lines represent the 95% confidence intervals calculated by importance sampling. The red, dashed, vertical lines represent the 95% confidence intervals calculated by approximate profile likelihood. Black points show importance samples. Only parameter sets within 10 log-likelihood units from the maximum likelihood are shown.

**Figure S20:**
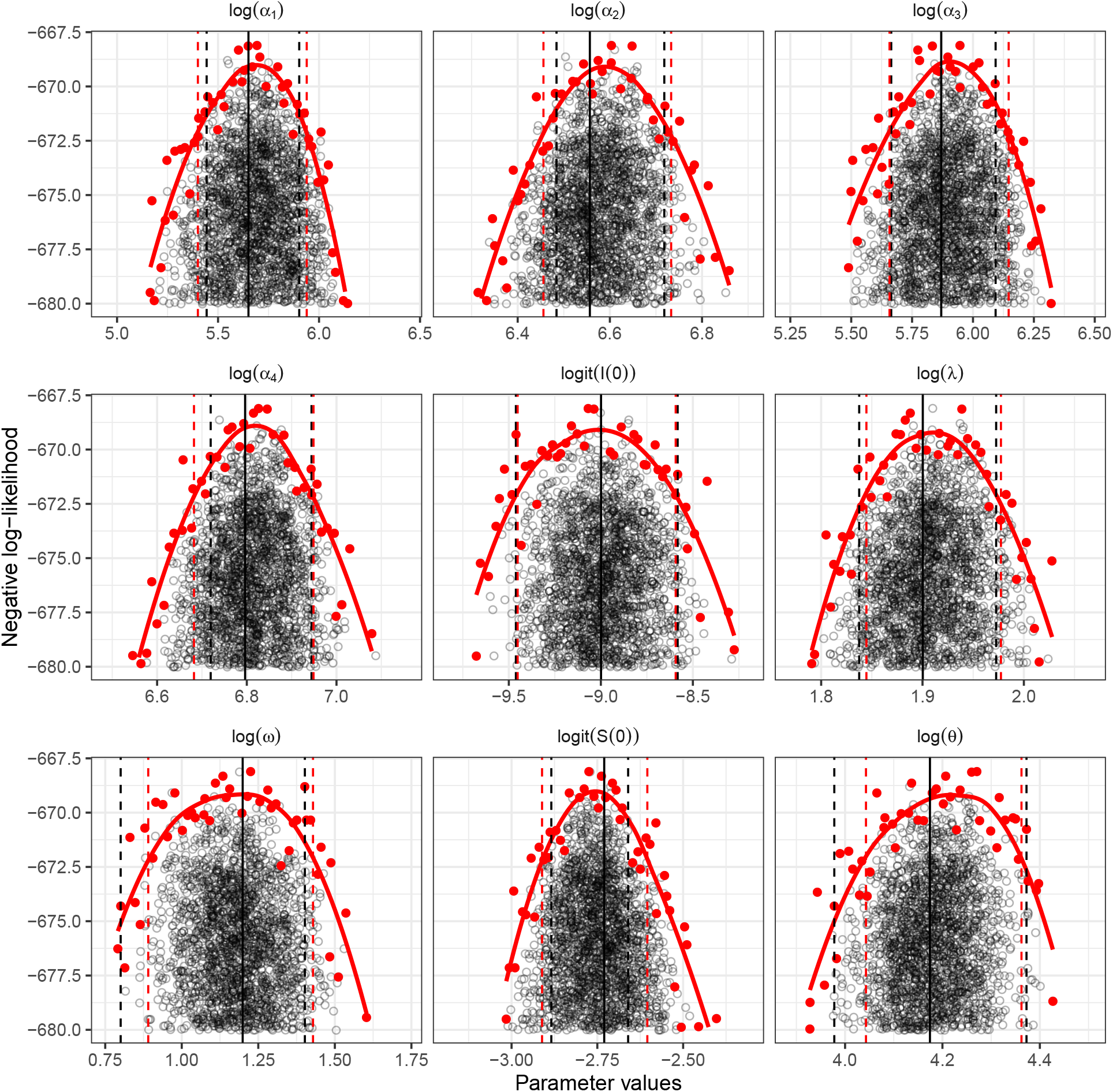
Parameter estimates and confidence intervals for New York, including the 2018 outbreak. The black, solid, vertical lines represent the MLEs. The black, dashed, vertical lines represent the 95% confidence intervals calculated by importance sampling. The red, dashed, vertical lines represent the 95% confidence intervals calculated by approximate profile likelihood. Black points show importance samples. Only parameter sets within 10 log-likelihood units from the maximum likelihood are shown.

**Figure S21:**
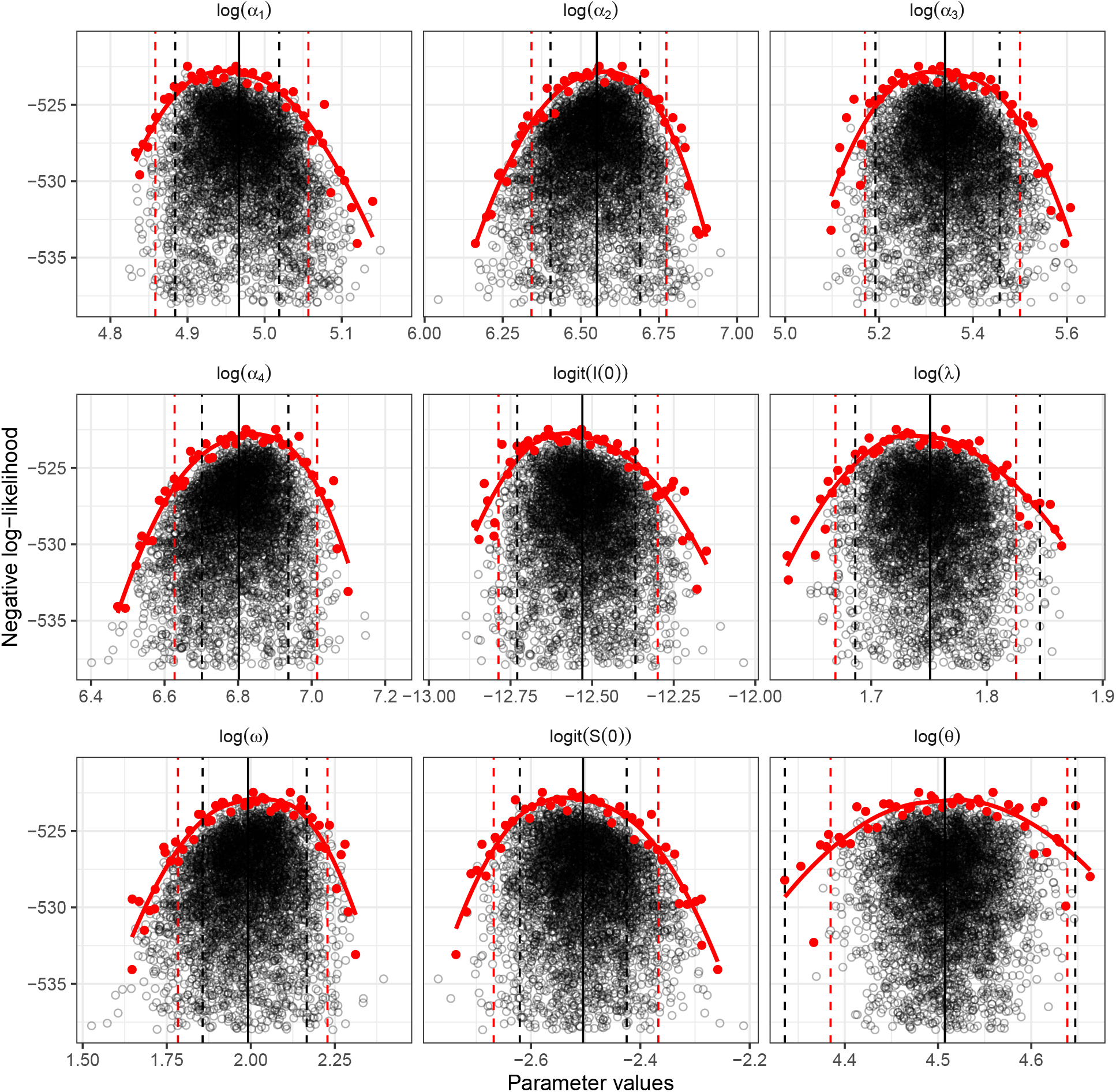
Parameter estimates and confidence intervals for Ohio, including the 2018 outbreak. The black, solid, vertical lines represent the MLEs. The black, dashed, vertical lines represent the 95% confidence intervals calculated by importance sampling. The red, dashed, vertical lines represent the 95% confidence intervals calculated by approximate profile likelihood. Black points show importance samples. Only parameter sets within 10 log-likelihood units from the maximum likelihood are shown.

**Figure S22:**
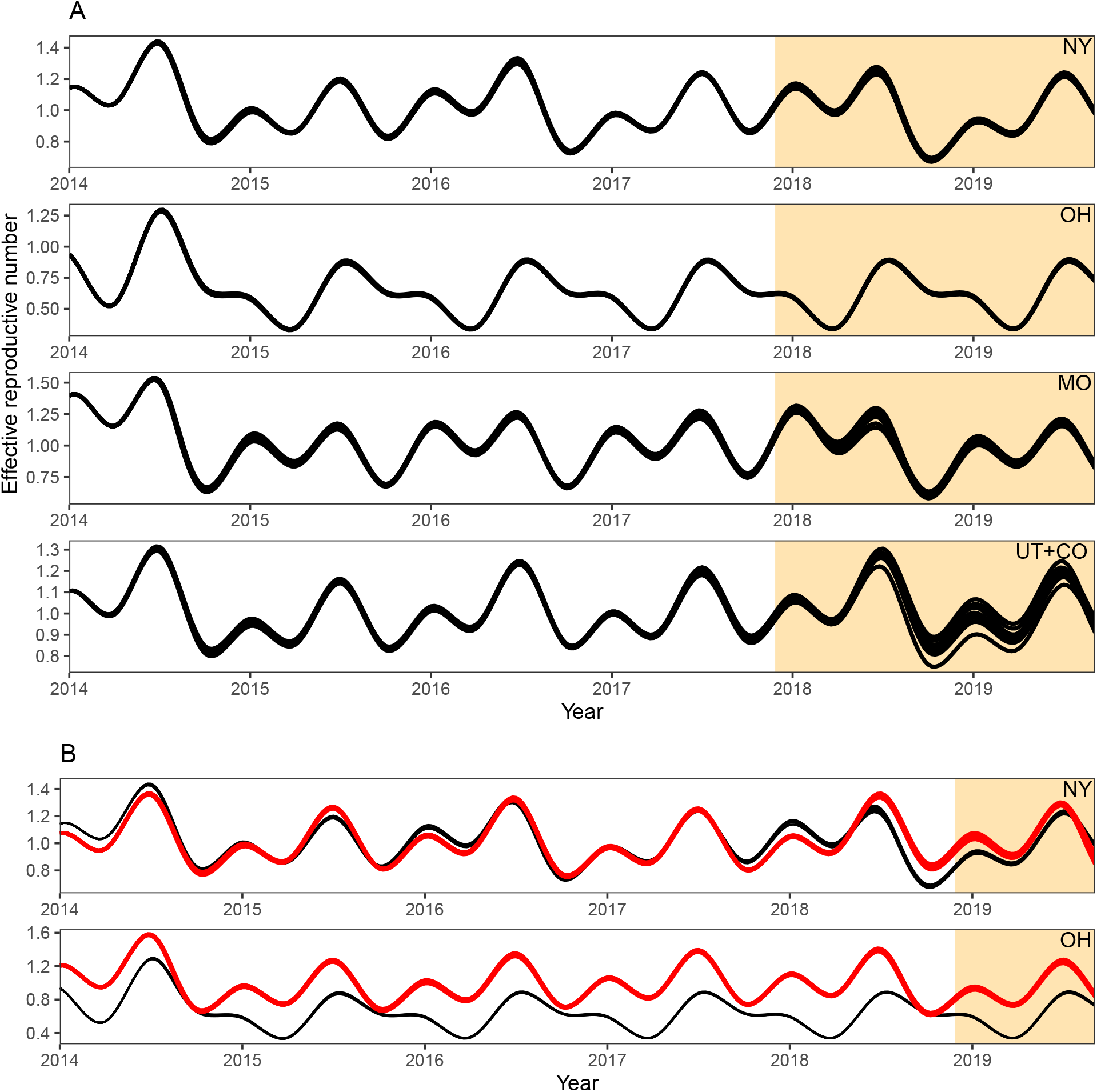
Estimates of the effective reproductive number over time. The effective reproductive number is calculated by multiplying the basic reproductive number *ℛ*_0_ with the proportion of susceptible individuals *S*_*t*_ over time. Predictions were made by simulating forward from 10 particle filtered trajectories.

**Table S1:** Population sizes in each state. Total population sizes represent population sizes in each region as of 2018. Adjusted population sizes represent the sum of population sizes of the cities from which EV/RV samples are collecte as of 2018.

| Region | Total population size | Adjusted population size |
| --- | --- | --- |
| New York | 19,542,209 | 8,766,726 |
| Ohio | 11,689,442 | 1,167,508 |
| Missouri | 6,126,452 | 794,756 |
| Utah+Colorado | 8,856,669 | 818,544 |

**Table S2:** Maximum likelihood estimates of the SIR model parameters for 2014– 2017 outbreaks. Spline coefficients *α*_*i*_ have a unit of year^*−*1^.

| Region | $\alpha_1$ | $\alpha_2$ | $\alpha_3$ | $\alpha_4$ | $\omega$ | $\lambda$ | $S(0)$ | $I(0)$ | $\theta$ |
| --- | --- | --- | --- | --- | --- | --- | --- | --- | --- |
| New York | 523 | 1253 | 647 | 1459 | 1.14 | 7.29 | 0.0363 | 3.48E-05 | 85 |
| Ohio | 280 | 445 | 22 | 663 | 202 | 4.14 | 0.0891 | 1.51E-05 | 179 |
| Missouri | 203 | 1023 | 411 | 1071 | 1.54 | 5.35 | 0.0584 | 3.44E-4 | 317 |
| Utah+Colorado | 304 | 580 | 343 | 699 | 0.08 | 4.67 | 0.0735 | 1.97E-4 | 505 |

**Table S3:** Maximum likelihood estimates of the SIR model parameters for 2014– 2018 outbreaks. Spline coefficients *α*_*i*_ have a unit of year^*−*1^.

| Region | $\alpha_1$ | $\alpha_2$ | $\alpha_3$ | $\alpha_4$ | $\omega$ | $\lambda$ | $S(0)$ | $I(0)$ | $\theta$ |
| --- | --- | --- | --- | --- | --- | --- | --- | --- | --- |
| New York | 284.47 | 703.87 | 354.10 | 894.45 | 3.32 | 6.67 | 0.0612 | 1.24E-04 | 65 |
| Ohio | 143.57 | 700.13 | 208.58 | 899.89 | 7.33 | 5.76 | 0.0755 | 3.62E-06 | 91 |

**Table S4:** 95% confidence intervals of the SIR model parameters for 2014–2017 outbreaks. Confidence intervals are calculated via importance sampling. Spline coefficients *α*_*i*_ have a unit of year^*−*1^.

| Region | $\alpha_1$ | $\alpha_2$ | $\alpha_3$ | $\alpha_4$ | $\omega$ | $\lambda$ | $S(0)$ | $I(0)$ | $\theta$ |
| --- | --- | --- | --- | --- | --- | --- | --- | --- | --- |
| New York | 457–625 | 1136–1389 | 517–773 | 1336–1590 | 0.86–1.52 | 6.77–7.95 | 0.0325–0.0401 | 2.43E-5–4.93E-5 | 60–104 |
| Ohio | 199–424 | 351–527 | 4–91 | 541–756 | 139.63–290.52 | 3.71–4.53 | 0.0761–0.112 | 4.533E-6–2.956E-5 | 110–331 |
| Missouri | 140–293 | 847–1367 | 322–550 | 914–1438 | 1.17–1.88 | 4.98–6.06 | 0.0442–0.0697 | 2.41E-6–5.26E-6 | 169–786 |
| Utah+Colorado | 212–416 | 428–768 | 225–450 | 543–894 | 0.03–0.17 | 4.26–5.20 | 0.0579–0.0955 | 1.12E-4–2.90E-4 | 115–3473 |

**Table S5:** 95% confidence intervals of the SIR model parameters for 2014–2018 outbreaks. Confidence intervals are calculated via importance sampling. Spline coefficients *α*_*i*_ have a unit of year^*−*1^.

| Region | $\alpha_1$ | $\alpha_2$ | $\alpha_3$ | $\alpha_4$ | $\omega$ | $\lambda$ | $S(0)$ | $I(0)$ | $\theta$ |
| --- | --- | --- | --- | --- | --- | --- | --- | --- | --- |
| New York | 231–366 | 655–827 | 288–443 | 828–1037 | 2.22–4.06 | 6.28–7.19 | 0.053–0.065 | 7.78E-5–1.87E-4 | 53–79 |
| Ohio | 132–151 | 604–804 | 180–234 | 814–1029 | 6.40–8.73 | 5.40–6.33 | 0.068–0.081 | 2.96E-6–4.26E-6 | 76–104 |

## Ethical statement

The data are de-identified at the institutions prior to export to the Syndromic Trends database according to HIPAA standards and BioFire enters into a Data Use Agreement with each institution which controls the use of the data and protects against re-identification of any individual. This analysis relied on de-identified data only. We did not seek for additional approval to conduct this study from an ethical review board.

## Notes

### Competing Interest Statement

LM is a former employee of bioMérieux, Inc, or its subsidiaries. bioMérieux markets the BioFire System and Syndromic Trends.

### Summary of Updates

Competing Interest Statement updated.

